# A Multi-City COVID-19 Categorical Forecasting Model Utilizing Wastewater-Based Epidemiology

**DOI:** 10.1101/2024.09.16.24313752

**Authors:** Naomi Rankin, Samee Saiyed, Hongru Du, Lauren Gardner

**Author notes:** Corresponding author. *Email address:* (Naomi Rankin).

## Abstract

The COVID-19 pandemic highlighted shortcomings in forecasting models, such as unreliable inputs/outputs and poor performance at critical points. As COVID-19 remains a threat, it is imperative to improve current forecasting approaches by incorporating reliable data and alternative forecasting targets to better inform decision-makers.

Wastewater-based epidemiology (WBE) has emerged as a viable method to track COVID-19 transmission, offering a more reliable metric than reported cases for forecasting critical outcomes like hospitalizations. Recognizing the natural alignment of wastewater systems with city structures, ideal for leveraging WBE data, this study introduces a multi-city, wastewater-based forecasting model to categorically predict COVID-19 hospitalizations.

Using hospitalization and COVID-19 wastewater data for six US cities, accompanied by other epidemiological variables, we develop a Generalized Additive Model (GAM) to generate two categorization types. The Hospitalization Capacity Risk Categorization (HCR) predicts the burden on the healthcare system based on the number of available hospital beds in a city. The Hospitalization Rate Trend (HRT) Categorization predicts the trajectory of this burden based on the growth rate of COVID-19 hospitalizations. Using these categorical thresholds, we create probabilistic forecasts to retrospectively predict the risk and trend category of six cities over a 20-month period for 1, 2, and 3 week forecasting windows.

We also propose a new methodology to measure forecasting model performance at change points, or time periods where sudden changes in outbreak dynamics occurred. We also explore the influence of wastewater as a predictor for hospitalizations, showing its inclusion positively impacts the model’s performance. With this categorical forecasting study, we are able to predict hospital capacity risk and disease trends in a novel and useful way, giving city decision-makers a new tool to predict COVID-19 hospitalizations.

## 1. Introduction

The COVID-19 pandemic had an unprecedented impact on public health and healthcare systems in the United States, resulting in more than 6.7 million hospitalizations and 1.1 million deaths as of February 3, 2024 (Centers for Disease Control and Prevention, 2024a). Various surveillance methods have been used to track the spread of the disease, including individual testing and surveys. However, these traditional approaches face challenges such as inconsistent sample sizes, which fluctuate over time, biases in individuals’ willingness to get tested or participate in surveys, and the underreporting of at-home test results (Nixon et al., 2022; Li et al., 2023; Reese et al., 2021; Rubin, 2021). In order to address these challenges, wastewater-based epidemiology (WBE) has emerged as a viable method to track virus spread in a community by measuring SARS-CoV-2 concentration trends in wastewater (Feng et al., 2021). WBE offers a non-invasive, aggregated, low-cost approach for monitoring COVID-19 infection trends and potential incidence in a given catchment area (Shah et al., 2022; Bibby et al., 2021). Several studies have found correlations between SARS-CoV-2 levels in wastewater and lagged COVID-19 cases, hospitalizations, and deaths (Shah et al., 2022; Galani et al., 2022; Kanchan et al., 2024). These findings suggest that WBE is a viable representation of community disease prevalence and has the potential to serve as a valuable input for COVID-19 forecasting models.

Recent studies have incorporated SARS-CoV-2 load in wastewater as a key variable to help forecast COVID-19 hospitalizations. A recent study forecasted weekly COVID-19 hospitalizations for 159 counties in the U.S. using wastewater data, demonstrating that forecasting models that incorporate WBE can be effective at predicting county-level COVID-19 hospital admissions 1 to 4 weeks in advance (Li et al., 2023). Hill et al., incorporated WBE into a generalized linear mixed model to predict COVID-19 hospitalizations at 4 different geographic scales (sewershed level, county level, regional level, and state level) for 56 counties in New York State to demonstrate the higher predictive power provided by incorporating wastewater into forecasting models (Hill et al., 2023). Many studies have also incorporated WBE into different modeling frameworks to predict COVID-19 hospitalizations outside of the United States. For example, Schenk et al., incorporated WBE in multivariate regression models to infer national hospitalization bed occupancy in Austria by aggregating SARS-CoV-2 load from local wastewater treatment plants to a national level. The study determined that, at a national level, increasing the number of monitored wastewater plants provides more accurate forecasts. (Schenk et al., 2023). Zamarreño et al., developed a dynamic artificial neural network to predict the number of hospitalized patients in Valladolid, Spain, utilizing wastewater-based epidemiology. The accuracy of the model was improved by forecasting a categorical risk level which aligns with warning levels established by the Regional Health Administration in Castile and Leoón (Zamarreño et al., 2024). Finally, Vaughan et al. incorporated WBE into a random forest algorithm to forecast COVID-19 hospitalizations in Scotland, Catalonia, Ohio, the Netherlands, and Switzerland. To improve model performance, the authors advocate for inputting data with a high sample frequency to ensure adequate training set size (Vaughan et al., 2023).

While these studies demonstrate the promise of WBE for forecasting COVID-19 hospitalizations, certain areas offer potential for further exploration and improvement. Firstly, most existing studies forecast continuous COVID-19 health outcomes as targets, such as the raw number of COVID-19 cases and hospitalizations. However, these continuous forecasts struggle to effectively communicate uncertainty to decision-makers, which can result in misinterpretation of model results and can be challenging to translate into actionable outcomes (Nixon et al., 2022). There is also a need to compare the efficacy of various non-continuous forecasts, such as those based on resource availability or previous week trends, to ensure that the result is accurate and clear. Secondly, few models are built at the city level, which aligns with wastewater testing catchment areas due to the presence of centralized wastewater treatment facilities that offer the best support for local-level decision making (Sen et al., 2023; Hillary et al., 2020). Thirdly, the potential of wastewater data to enhance model performance during critical periods, particularly those characterized by rapid or sudden trends, remains uncertain. Therefore, model evaluations should focus specifically on these periods to better understand and evaluate the model’s utility.

In this paper, we expand on the current literature by presenting an interpretable categorical short-term forecasting model using Generalized Additive Models (GAMs) that incorporates WBE data to predict weekly COVID-19 hospitalizations for 6 cities in the United States from January 2021 to November 2022. Our multi-city model generates two actionable categorical outputs for city-level decision-makers: Hospitalization Capacity Risk (HCR), which predicts the strain on hospital resources based on available beds, and Hospitalization Rate Trend (HRT), which forecasts the future trajectory of disease transmission based on hospitalization trends. This model generates 1-week, 2-week, and 3-week ahead forecasts by incorporating SARS-CoV-2 wastewater load, recent hospitalization rates, vaccination rates, prior infection data, and static variables like the COVID-19 Community Vulnerability Index. We assessed model performance for both categorical outputs across six cities for over 80 weeks, utilizing five distinct error metrics. To enhance our understanding of the model’s accuracy and utility, we examined performance at change points, which are defined as any week where the true hospitalization category is different than the previous week. The GAM framework allows for the evaluation of variable contribution, highlighting the value of using WBE data for forecasting COVID-19 health outcomes at the city level. The proposed model offers a valuable tool for decision-making support due to its simple design, reliance on readily available data, and production of easily interpretable and reliable forecasts.

## 2. Material and Methods

In this section, we present the design of our study, beginning with an overview of the data collection and preprocessing procedures in Section 2.1, as well as the design of the target variables in Section 2.2. A summary table of all variables is provided in Table 3. We then introduce the GAMs utilized in this study in Section 2.3 and discuss the error metrics used to evaluate the model’s performance in Section 2.4.1. Furthermore, we propose a novel approach for measuring forecasting capabilities at critical points in Section 2.4.2.

### 2.1 Input Variables

This study utilizes SARS-CoV-2 wastewater surveillance data alongside other epidemiological metrics such as hospitalizations, previous infections, vaccination coverage, and local vulnerability indicators to forecast weekly COVID-19 hospitalizations for six U.S. cities. City boundaries were delineated using USPS ZIP codes identified through the USPS ZIP code lookup tool (United States Postal Service, 2024). Detailed descriptions of each input variable are provided below.

#### Hospitalizations (H)

In this study, we utilize COVID-19 hospitalization data as both an input to our model and to define our categorical targets, the latter of which is presented in Section 2.2. We obtained raw hospitalization data from the U.S. Department of Health and Human Services (HHS) COVID-19 Reported Patient Impact and Hospital Capacity by Facility dataset (United States Government Department of Health & Human Services, 2024). For each city *i*, the weekly reported hospitalizations at week 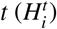 were summed across all facilities *f* within the zip codes belonging to the city as follows:

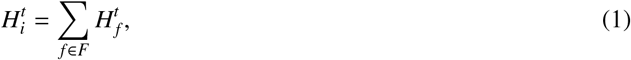

where *f* is a healthcare facility within the set of all facilities *F* in the zip codes of a the city at week *t*. The 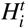were then logged to generate the input variable for the model to normalize for city-specific hospitalization scales. The detailed data cleaning procedures of the 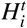 are described in **Supplementary A**. Previous COVID-19 hospitalizations are considered indicators of future trends, (Hill et al., 2023) thus at each week *t*, we include the hospitalization data from the previous three weeks as separate input variables 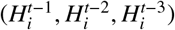. This approach enhances the model’s predictive capabilities by incorporating recent hospitalization dynamics.

#### SARS-CoV-2 Wastewater Viral Load (WW)

Given that SARS-CoV-2 concentrations in wastewater are leading indicators of COVID-19 hospitalizations, we included city-level SARS-CoV-2 wastewater viral loads as an input variable to forecast city-level hospitalizations. The SARS-CoV-2 load in wastewater was obtained from state and city dashboards (see detailed reference in Table 1). For each city, we include SARS-CoV-2 concentrations for all sewersheds partially or completely within the city boundaries.

**Table 1:** Cities of interest in the study and their wastewater collection systems and dates.

| City | Population | Number of Treatment Plants | Sampling Frequency | Source |
| --- | --- | --- | --- | --- |
| Charlotte, NC | 942,437 | 3 | Every 3 days | (North Carolina Department of Health and Human Services, 2024) |
| Denver, CO | 1,399,707 | 1 | Daily | (Colorado Department of Public Health and Environment, 2024) |
| Houston, TX | 3,206,416 | 39 | Daily | (Kinder Institute Urban Data Platform, 2024) |
| New York, NY | 8,570,761 | 14 | Thrice Weekly | (New York City Department of Health and Mental Hygiene, 2024) |
| San Diego, CA | 1,387,376 | 1 | Every 2 days | (Lab, 2024) |
| San Francisco, CA | 865,933 | 2 | Daily | (California State Water Resources Control Board, 2024) |

The SARS-CoV-2 wastewater viral loads were reported as viral gene copies/L at varying frequencies across cities. All viral load data was aggregated to a one-week reporting frequency and logged to normalize the input variable across cities. We evaluated the preceding relationship between wastewater viral loads and COVID-19 hospitalization rates by identifying the leading weeks where there was a Pearson correlation coefficient of at least 0.75 between the two. A detailed description of these correlations can be found in Figure S2 in **Supplementary A**. Accordingly, each week, we utilize the wastewater viral loads from the previous three weeks as separate input variables.

#### Past Infections (PI)

Natural immunity has been shown to confer significant protection against COVID-19 reinfection and severe outcomes (Pooley et al., 2023). To capture the impact of natural immunity, we adapted the past infection metric (PI) from a prior study as a proxy for population natural immunity (Du et al., 2024a). This variable quantifies the number of reported COVID-19 infections within the past three months, using daily reported county level case data from the Johns Hopkins COVID-19 Dashboard (Dong et al., 2020) aggregated to a weekly level. Due to the lack of consolidated reported case data at the city level, we use the data from the county that encompasses the majority of the city as a proxy for city-level case counts. The PI formulation is as follows:

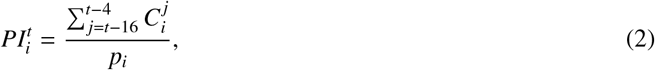

where 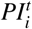 denotes the total reported infections in county *i* during the previous 12 weeks, 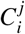 is the number of reported cases for city *i* at week *j*, and *p*_*i*_ is the population for city *i*.

#### Full Vaccination Coverage (VC)

In addition to the incorporation of PI as a proxy for natural immunity, we included the cumulative percentage of the population that is fully vaccinated to account for the impact of vaccine-induced immunity on COVID-19 hospitalizations. We obtained weekly COVID-19 vaccination data from the Immunization Information System at the Centers for Disease Control and Prevention (Centers for Disease Control and Prevention, 2023). Similar to the reported case data, this national vaccination dataset tracks vaccine uptake at the county level. Therefore, we mapped city vaccination coverage data from the most representative county. Fully vaccinated is defined as the total number of individuals who have completed a primary vaccine series, either recieving the second dose of a two-dose series or one dose of a single-dose series. The percentage of fully vaccinated is calculated by dividing the fully vaccinated population by the total county population.

#### COVID-19 Community Vulnerability Index (CCVI)

In order to account for population-level susceptibility to adverse disease outcomes, we utilize the COVID-19 Community Vulnerability Index (CCVI), which was developed by the Surgo Foundation (Ventures, 2021). This index is an adaptation of the CDC Social Vulnerability Index with a focus on the specific risk factors of COVID-19 for all zip codes in the United States. The CCVI covers seven themes (Socioeconomic Status, Minority Status and Language, Housing Type, and Transport, Epidemiological Factors, Healthcare System, High Risk Environments, and Population Density), giving each zip code a numerical value on a scale from 0 to 1, where a 1 indicates a community has a high vulnerability in that theme. For each of the cities in the study, we quantify the vulnerability themes by aggregating the zip-code level CCVI values as follows:

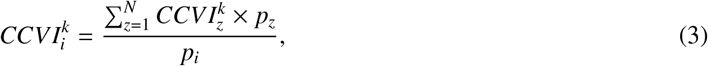

where *N* is the set of all zip codes *z* within a city *i*, and *p* is the population. The zip code-level CCVI values were aggregated to the city level by weighting each by its zip code population and then normalizing by the total city population to ensure the variable ranges between 0 and 1. We repeat this aggregation for each of the seven themes *k* and use them as static inputs for our model to represent local vulnerability to adverse outcomes of COVID-19 throughout the study period.

### 2.2 Target Design: Hospitalization Categorization

This study aims to provide city-level decision-makers with interpretable and actionable forecasts for COVID-19 hospitalizations by enhancing robustness against reporting issues. Rather than relying on traditional numerical predictions of hospitalization counts, we forecast risk categories that are derived from the rate of COVID-19 hospitalizations per 100,000 people. We propose two distinct categorization models, each emphasizing different aspects of the future impact of COVID-19 on a city healthcare system: the Hospitalization Capacity Risk (HCR) Categorization, which predicts the burden on the healthcare system and the Hospitalization Rate Trend (HRT) Categorization, which predicts the trajectory of COVID-19 hospitalization trends.

#### Hospitalization Capacity Risk (HCR) Categorization

Hospital demand can be used to measure times of overburdened healthcare systems, indicating when personal risk of infection is at its highest. We develop the HCR as a 5-tier categorization model based on hospital demand, defined as the static ratio between observed hospital admission rate and average number of available hospital beds for COVID-19 forecasted in a city. The HCR categorization is defined as follows:

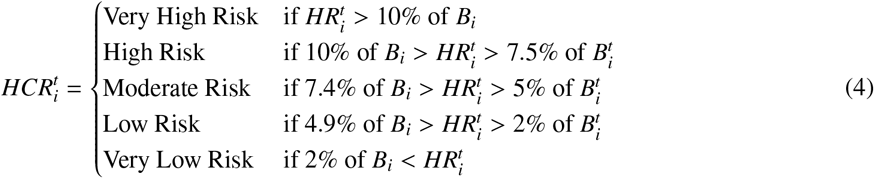

Where 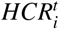 is the Hospitalization Capacity Risk category for city *i* at week *t*, 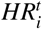 is the forecasted COVID-19 hospitalization rate per 100,000 people for city *i* at week *t*, and *B*_*i*_ is the average number of available hospital beds per 100,000 people for city *i* across the study period. The thresholds of this categorization reflect the static capacities of each specific city’s healthcare system.

The thresholds are designed after the City of Austin stage alert system (Yang et al., 2021). They created a static 4-tier alert system based on the percent of filled ICU beds and the 7-day average of COVID-19 hospitalizations at a city level from March 2020 to September 2021. We calculate the percentage of utilized COVID-19 beds for Austin equivalent to the threshold number of hospitalizations determining each stage threshold. Additionally, we introduce a fifth category of highest risk to further specify the intensity of burden during times of peak hospitalizations. This categorization provides a more detailed and city-specific understanding of risk than the CDC 3-tier risk metric that identifies high, medium, and low hospital admission rates per 100,000 population based on a single threshold for all US locations (Centers for Disease Control and Prevention, 2024b). By increasing the number of high-risk categories, we expand on the CDC 3-tier community risk system and the Austin 4-tier system while still maintaining thresholds that reflect predetermined foundations for risk levels.

Figure 1 illustrates the five risk categories of the HCR for the six cities, where the black line indicates the observed hospitalization rate per 100,000 population, and the colored bars indicate the risk category. Across cities, there are common times of higher risk, such as January 2022 during the peak of the Omicron wave, but the city-specific dynamics vary by risk level at other times. Table 2 presents the distribution of categories for the HCR across the study period and all locations. The majority of weeks are assigned as Low and Very Low risk, indicating that times of very high community risk are less prevalent across multiple locations over the study period.

**Table 2:** Thresholds for Hospitalization Rate Trend Categorization and distribution of categories.

|  | Category | Threshold | Percent of Study Period |
| --- | --- | --- | --- |
| HCR | Very High Risk | >10% | 14.2% |
|  | High Risk | 7.5 to 10% | 8.8% |
|  | Moderate Risk | 5 to 7.4% | 16.6% |
|  | Low Risk | 2 to 4.9% | 31.9% |
|  | Very Low Risk | <2% | 28.5% |
| HRT | Large Increase | >20% | 22.5% |
|  | Increase | 10 to 19.9% | 12.1% |
|  | Stable | -10 to 10% | 19.8% |
|  | Decrease | -10 to -19.9% | 17.1% |
|  | Large Decrease | <-20% | 28.5% |

**Table 3:** Inputs and Targets of the model.

| Variable | Calculation/Collection | Units | Type | Usage |
| --- | --- | --- | --- | --- |
| SARS-CoV-2 Load in Wastewater | State/City Dashboards (3 preceding weeks) | log(N gene copies/L) | Dynamic | Input |
| Past Infections | HHS | Percent | Dynamic | Input |
| Percentage Vaccinated | IIS | Percent | Dynamic | Input |
| CCVI | Surgo Ventures | Ratio | Static | Input |
| Log Hospitalization Rate | HHS (3 preceding weeks) | log(hospitalizations/100k people) | Dynamic | Input |
| Hospitalization Capacity Risk Categorization | HCR Methodology | Risk Categories from 1 to 5 | Static | Target |
| Hospitalization Rate Trend Categorization | HRT Methodology | Trend Categories from -2 to 2 | Dynamic | Target |

**Figure 1.**
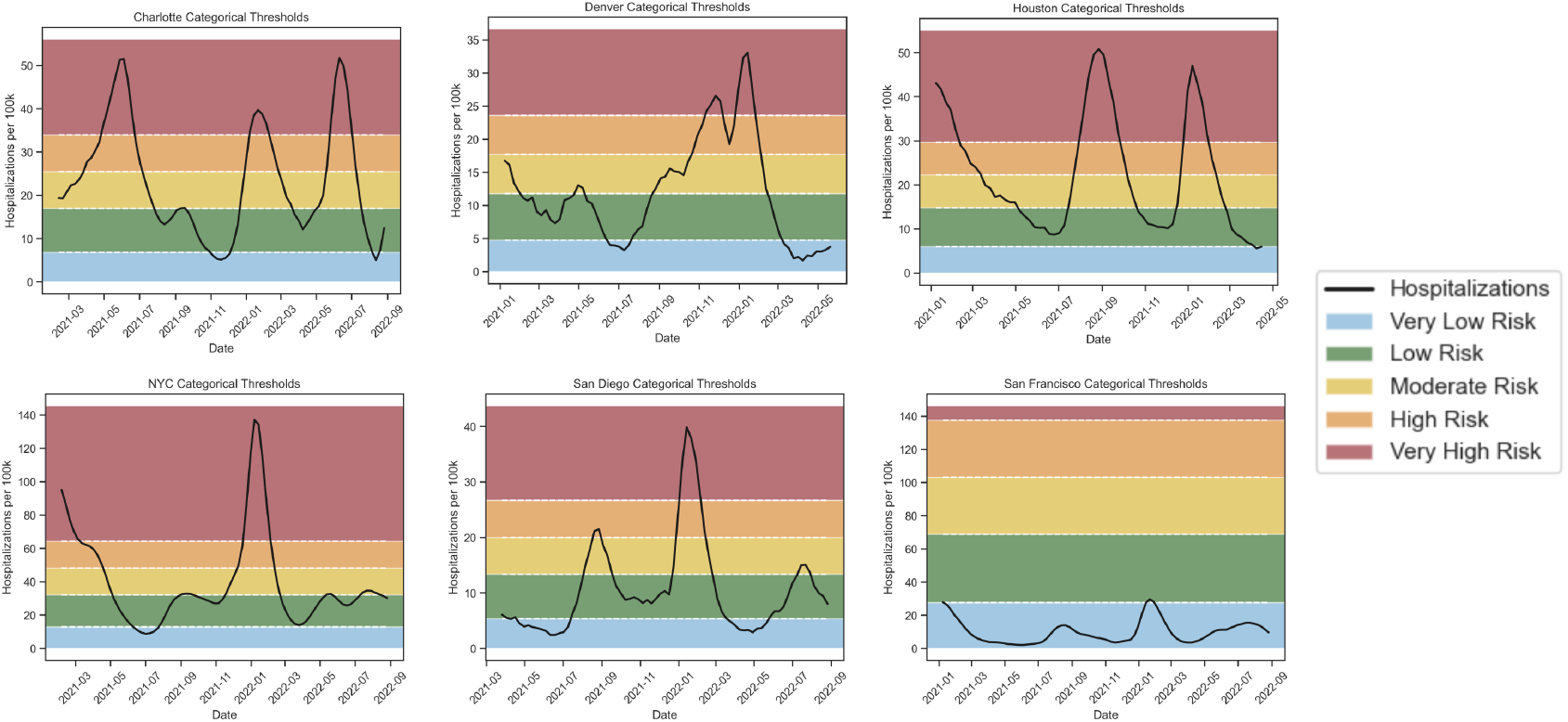
Hospitalization Capacity Risk Categorization Visualization. HCR category thresholds by color in Charlotte, NC; Denver, CO; Houston, TX; New York, NY; San Diego, CA; and San Francisco, CA. The background color indicates the risk category. The dotted white lines indicate the thresholds, and the black line is the true hospitalization rate.

#### Hospitalization Rate Trend (HRT) Categorization

Observing trends in the rate of change of COVID-19 hospitalizations provides public health decision makers with a dynamic understanding of the change in burden on a city’s healthcare system. We develop the HRT categorization to capture city-level COVID-19 hospitalization trends to support broader city-level decision-making to describe how quickly the number of hospitalizations is changing between weeks, providing a complementary metric to the static categorization of the HCR. This categorization is also derived from the weekly average COVID-19 hospitalization rate per 100,000 people (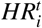). This metric is useful to forecast periods of large changes for public planning purposes and is inspired by the FluSight forecasting target (FluSight-forecast-hub, 2024). We define the rate trend 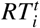 as the change in COVID-19 hospitalizations for a city *i* in a given week *t* relative to the average change of the prior three weeks. The rate change is calculated as:

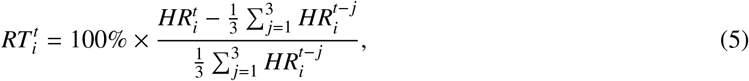

where 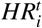 is the hospitalizations per 100,00 at week *t* for city *i*, and the summation is the mean hospitalization rate for the previous 3 weeks in the same city. Using this growth rate, we define the HRT categorization as:

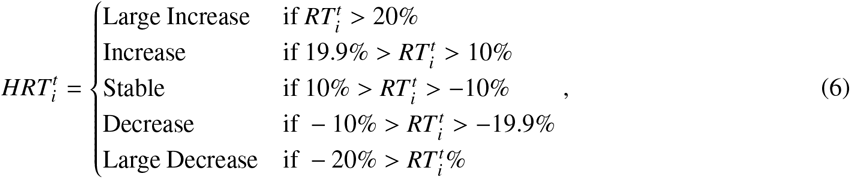

The thresholds of this categorization reflect the changing dynamics of the COVID-19 hospitalization trends.

Figure 2 illustrates the rate trend categorical assignment for all six cities. The black line indicates the change in hospitalizations compared to previous weeks. The rate trend differs between cities based on their individual dynamics, not an aggregated national trend, providing more specific and actionable information. Table 2 presents the distribution of categories for the HRT across the study period for all locations. Over our study period, most cities for most weeks are in the Large Increase or Large Decrease category, indicating the volatile trends of COVID-19 that have been historically difficult to predict.

**Figure 2.**
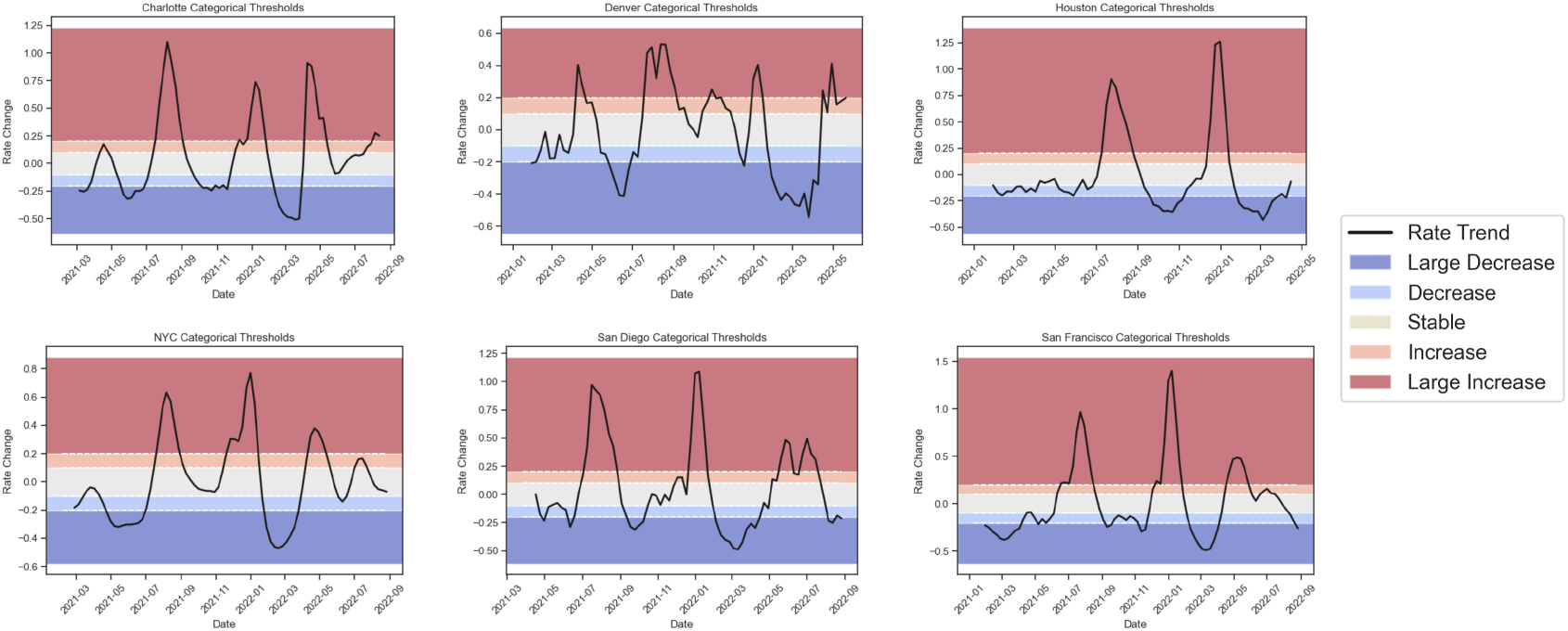
Hospitalization Rate Trend Categorization Visualization. HRT category thresholds by color in Charlotte, NC; Denver, CO; Houston, TX; New York, NY; San Diego, CA; and San Francisco, CA. The dotted white lines indicate the thresholds, the black line represents the observed rate change, and the background color indicates the category.

### 2.3 Model

We develop 1, 2, and 3-week out forecasts of weekly COVID-19 hospitalizations for the 82-week period from January 2021 to September 2022 using wastewater viral load, past infections, the full vaccination coverage, previous hospitalizations, and 7 social vulnerability indices, summarized in Table 3. These forecasts are created via Generalized Additive Models (GAMs), an additive model that can learn non-linear predictor variables by modeling the outcome as a sum of spline functions of each predictor. We fit a distinct GAM for each forecasting window (1-week, 2-week, and 3-week), each utilizing the same model formulation:

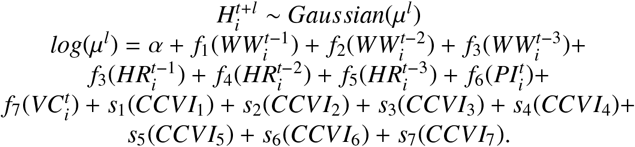

Where *l* is the forecasting window, taking values of 1, 2, or 3. Each *f*_*i*_ coefficient indicates the spline smooth function of the dynamic variables, *α* indicates the intercept and *µ*^*l*^ is the mean of the hospitalization distribution *l* weeks ahead. Each *s*_*i*_ represents the parametric coefficient of the static variables. In order to mimic real-time forecasting practices, we used an expanding training window. This method involves initially training the model on a small set of data and then gradually expanding the training set as new data becomes available. In this study, the model is first trained on data before February 26, 2021. As each new week of data is collected, it is added to the training set, thereby expanding the window of historical data used for model training. This analysis was done using the pyGAM package in Python 3.9.13.

The continuous predictions are then converted into discrete categories to generate categorical predictions for each forecasting window. To create a probabilistic distribution over these categories, we performed prediction interval sampling to generate multiple continuous outcomes. We then compute the frequency with which these interval sample falls into each category. Specifically, for each week and city, we utilize 100 *H*^*t*^ values representing the upper and lower bounds of 1% to 99% prediction intervals. Each interval value is then converted to a categorical label based on the procedure described in Section 2.2. The final forecast for each week and city is the most frequent label among 100 probabilistic samples, selecting the categorical prediction with the greatest overall agreement.

### 2.4 Evaluation

This section details the various ways in which we measure forecasting performance. We define and justify the error metrics used to determine categorical forecast performance, as well as describe a novel procedure to determine model accuracy at the critical points where the disease burden is increasing or decreasing.

#### 2.4.1 Error Metrics

We evaluate our model using five error metrics: 1) Accuracy, 2) Mean Square Error (MSE), 3) Weighted Mean Square Error (WMSE), 4) Brier Score, and 5) Brier Skill Score, each of which is established metrics for measuring categorical forecasting error (Bradley et al., 2008; Du et al., 2024b).

Accuracy measures the percentage of predicted labels that match the true labels. Although widely used as a baseline for evaluating model performance, accuracy alone does not convey how far off the predictions are from the true values. To address this, we use the MSE, which reflects the magnitude of error, with smaller values indicating better performance. However, MSE does not account for the uncertainty in the predictions. To incorporate uncertainty, we also use the WMSE, where the weights are the probabilities of each category. This metric penalizes the confidence in inaccurate forecasts, where a smaller value indicates a better performance. We also measure our inaccuracies with the Brier Score, an error metric designed specifically to measure the accuracy of probabilistic forecasts. It is calculated as the mean square difference between the predicted probability of each category to the actual probability of each category. A BS value lies between 0 and 1, where a 0 reflects perfect accuracy. From the Brier Score, we derive the Brier Skill Score, which compares our model performance to a baseline of random guessing with uniform probabilities for each category. A BSS less than 0 indicates that our predictions perform worse than the baseline, equal to 0 indicates that the prediction is equivalent to the baseline, and greater than 0 indicates that our predictions perform better than the baseline. The equations for each of these error metrics can be found in **Supplementary A**. Employing all five of these error metrics enables a comprehensive evaluation of our model performance for both precision and confidence, specific to categorical predictions.

#### 2.4.2 Model Performance at Change Points

A significant issue with hospitalization forecasting models during the peak of the COVID-19 pandemic was the failure to accurately predict time periods where sudden changes in outbreak dynamics occurred (Lopez et al., 2024). The ability to forecast such periods of rapid fluctuation sudden increases or decreases in reported hospitalization rates is critical for effective decision-making. To address this, we developed a novel method to evaluate model performance during these periods of rapid change, referred to as “change points”. We define a change point for both the HCR and HRT categorization as any week (*t*) where the true hospitalization category is different than the category of the previous week (*t*_−1_). We further classify change points into two categories: “upward shift”, where the category is higher than that of the previous week, such as going from a Low Risk to a Moderate Risk for the HCR, or a “downward shift”, where the category is lower than the previous week, such as going from a Large Increase to Stable for HRT.

We then evaluate each model’s performance during periods of rapid changes by assessing how accurately each model captures the correct category over all change points, as well as at upward and downward shifts specifically. We evaluated performance using MSE, WMSE, Brier Score, and Brier Skill Score for all change points in the model, the detailed definitions of which are in Section 2.4.1.

## 3. Results

In this section, we describe the model performance using the error metrics described in Section 2.4.1. We then provide an in-depth analysis of the HCR and HRT categorization model performances for the 2-week forecasting window in Section 3.1. The equivalent results for the 1 and 3-week forecasting windows are provided in Figures S1-S4 in **Supplementary B**. In Section 3.2, we present results for the change point evaluation. Finally, in Section 3.3, we present the results regarding variable importance.

### 3.1 Model Performance

Using the error metrics described in Section 2.4.1, we demonstrate the performance of the HCR and HRT categorization models for the 1, 2, and 3-week forecasting windows in Table 4. The Accuracy and BSS are error metrics where higher values indicate better model performance, whereas lower MSE, WMSE, and BS values indicate better model performance. The model performance reveals several key findings:

**Table 4:** A summary of model performance across various error metrics. ↑*/*↓ denotes if a higher/lower metric value signifies better performance.

|  |  | Accuracy ↑ | MSE ↓ | WMSE ↓ | BS ↓ | BSS ↑ |
| --- | --- | --- | --- | --- | --- | --- |
| HCR | 1-week | 88.8% | 0.114 | 0.106 | 0.157 | 0.885 |
|  | 2-week | 82.3% | 0.205 | 0.181 | 0.262 | 0.822 |
|  | 3-week | 74.8% | 0.382 | 0.269 | 0.341 | 0.748 |
| HRT | 1-week | 69.6% | 0.494 | 0.458 | 0.406 | 0.493 |
|  | 2-week | 56.4% | 1.24 | 1.044 | 0.566 | 0.292 |
|  | 3-week | 45.2% | 2.610 | 1.704 | 0.715 | 0.107 |

Both categorization models significantly outperform random guesses. A random guess model would yield an accuracy of 20% and a BSS of 0, whereas the HCR accuracy spans from 75 to 89% and the HRT accuracy spans from 46 to 70% across all forecasting windows. Notably, all BSS values remain above 0, further underscoring the predictive power of the models.

For both the HRT and HCR categorization models, performance worsens as the forecasting window increases, demonstrated by the decrease in accuracy and BSS and the increase in MSE, BS, and WMSE. The decrease in model performance over increasing forecasting windows mirrors the decrease in correlations between wastewater and future hospitalizations over time. This diminishing relationship reflects rapidly changing COVID-19 dynamics in a community which makes the further future more difficult to predict.

Across all error metrics, model performance when predicting capacity (HCR) is consistently better than when predicting trends (HRT). The fluctuating shifts in COVID-19 hospitalization trends tend to be more volatile than changes in capacity, making the HRT more difficult to accurately predict. We detail these specific categorization model performances in the following sections.

#### 3.1.1 Hospitalization Capacity Risk Categorization Results

In this section, we provide a detailed description of the performance of the HCR categorization model for the 2-week forecasting window. The 1 and 3-week forecast results are provided in Figures S1 and S2 of **Supplementary B**. In Figure 3, we demonstrate the overall categorical assignment accuracy, as well as the city specific categorical assignments over the study period.

**Figure 3.**
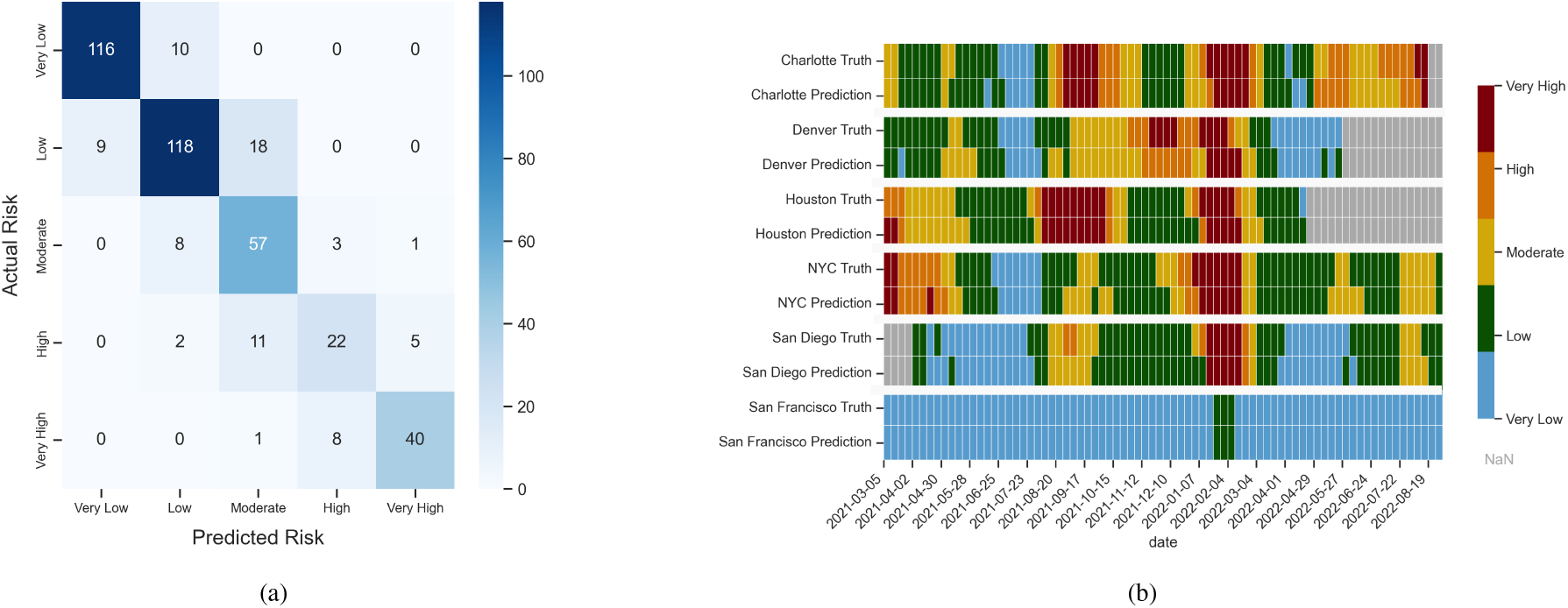
Summary of Model performance of Hospitalization Capacity Risk (HCR) Categorization, January 2021-September 2022. (a) Confusion Matrix detailing the accuracy of categorization for the HCR. Darker colors indicate that more assigned labels match the true labels for that category. (b) Hospital Capacity Risk true and predicted label for 2-week forecasts by city for January 2021 to September 2022. The color indicates the predicted category. Grey squares indicate that a prediction was not made for that week due to a lack of available data.

The HCR model performance can be described with a confusion matrix, which visualizes the accuracy of the HCR model across all cities and the entire study period for the 2-week forecast. In the matrix, each row represents the true categorical label of a week, and each column is the predicted label of that week. Each matrix entry contains the number of weeks where the true label is categorized as that predicted label, with incorrect categorical assignments are indicated by dissimilar row and column labels. In the confusion matrix in Figure 3a, the concentration of values along the diagonal indicates that the model accurately labels most forecasts. Off-diagonal values are primarily adjacent to the correct category, indicating that even when the model is incorrect, the magnitude of inaccuracy is relatively small.

Figure 3b compares the weekly HCR predictions to the true HCR categorizations for each city at a 2-week forecasting window, where the first row of the city illustrates the true weekly category and the second row illustrates the predicted category. The color indicates the categorical assignment, with grey indicating that predictions were not made due to a lack of available data over the study period from January 2021 to September 2022. Although the HCR is dependent on specific city dynamics, there are similar trends across all cities. There is a similarly high risk for all cities in January 2022, during the Omicron wave. San Francisco maintains a relatively low risk throughout the study period due to the low rate of hospitalizations per 100,000 people, but the subsequent increase at this time is still present. When the model predictions do not match the true category, it tends to predict the correct category with a minor time lag.

#### 3.1.2 Hospitalization Rate Trend Categorization Results

In this section, we focus on the performance of the HRT categorization model at the 2-week forecasting window. The 1 and 3-week forecast results are provided in Figures S3 and S4 in **Supplementary B**. In Figure 4, we demonstrate the overall categorical assignment accuracy with a confusion matrix, as well as the city specific categorical assignments with a time series plot equivalent to those shown in Figure 3.

**Figure 4.**
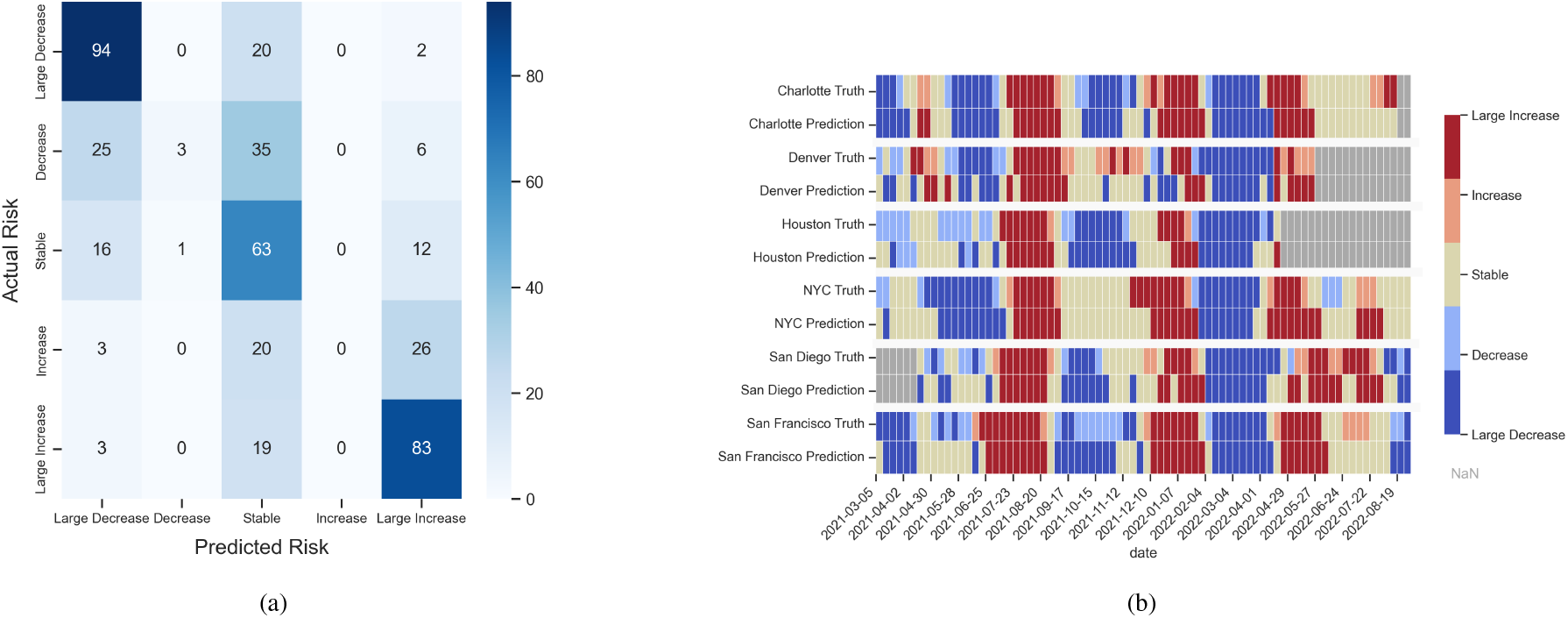
Summary of Model performance of Hospitalization Rate Trend (HRT) Categorization, January 2021-September 2022. (a) Confusion matrix detailing the accuracy of the categorization of the HRT. Darker colors indicate that more assigned labels match the true labels for that category. (b) City-specific Rate Trend target and 2-week prediction over time from January 2021 to September 2022. Grey squares indicate that a prediction was not made for that week due to a lack of available data.

The performance of the HRT categorization model for the 2-week forecasting window is illustrated in the confusion matrix in Figure 4a. The concentration of values at the top left and bottom right corners indicate that most COVID-19 hospitalization rate changes are large changes, and that the model can accurately predict Large Increase and Large Decrease growth rates. The concentration of values in the center of the confusion matrix indicates that weeks categorized as Stable, of which there are many, are also accurately predicted. The HRT model performance is worst during weeks categorized as Decrease and Increase, illustrated in their respective rows in the confusion matrix. In these rows, we can see that errors are nearly evenly split between overestimations and underestimations.

Figure 4b compares the weekly HRT predictions to the true HRT categorizations for each city at a 2-week forecasting window from January 2021 to September 2022 equivalently to Figure 3b. The top rows for each city illustrate the true HRT trends, revealing that periods of large increases are quickly followed by periods of large decreases, indicating the turbulent changes in COVID-19 hospitalizations over the study period. In the bottom row of each city we present the HRT predictions. Although the exact category is not always perfectly predicted, true increases generally yield forecasted increases and true decreases typically yield forecasted decreases. This phenomenon is expanded upon in Section 3.2.

### 3.2 Change Points Performance

In addition to building and evaluating interpretable city-level forecasting models, we also aim to develop a novel method to evaluate categorical forecasting model performance during periods of increases or decreases in reported hospitalization rates, denoted as change points. As outlined in the section 2.4.2, we measured model performance by determining model accuracy at all change points, as well as at upward and downward shifts specifically. We present the resulting WMSE scores in Table 5, and provide the MSE, Brier Score, and Brier Skill Score error metrics in Table T1 and T2 in **Supplementary B**. These results indicate that the HCR model consistently outperforms the HRT model, with the HCR predictions usually within a one category margin. Additionally, the error metrics generally worsen for farther ahead forecasts. Finally, we observe that both models perform worse at change points than in their overall evaluations.

**Table 5:** A summary of model performance using WMSE for change points for 1. all change points in the model, 2. from a lower to higher category (upward shift), 3. from a higher to lower category (downward shift). The final column 4. is model WMSE over all predictions (same as the WMSE column in Table 4). ↓ denotes that a lower error demonstrates better performance.

|  |  | Upward Shift | Downward Shift | All Change Points | Overall Model |
| --- | --- | --- | --- | --- | --- |
| HCR | 1 Week | 0.224 | 0.262 | 0.244 | 0.106 |
|  | 2 Week | 0.288 | 0.359 | 0.325 | 0.181 |
|  | 3 Week | 0.446 | 0.462 | 0.454 | 0.269 |
| HRT | 1 Week | 0.651 | 0.607 | 0.628 | 0.458 |
|  | 2 Week | 1.183 | 1.335 | 1.262 | 1.044 |
|  | 3 Week | 1.864 | 1.741 | 1.801 | 1.704 |

### 3.3 Variable Importance Analysis

This work aims to demonstrate the value of wastewater based epidemiology for improving disease surveillance capabilities. The following analysis illustrates the importance of each variable in improving forecast performance. We measure variable importance as the difference in mean average percent error (MAPE) of the continuous hospitalization forecasts when each variable is removed. This variable importance is measured for the 1, 2, and 3-week forecasts. This approach allows us to evaluate variable contribution independently of any categorical threshold selection biases. In Figure 5, we demonstrate the changes in MAPE after the removal of each variable, namely wastewater viral load, full vaccination coverage, past infections, and CCVI. For the 1 and 2-week forecasting window, the removal of wastewater leads to the highest increase in MAPE, indicating its critical role in improving forecasting performance. For the 3-week forecast, the removal of the CCVI leads to the largest increase in MAPE, indicating that in addition to wastewater, the community vulnerability indices serve an important role in understanding community risk and creating useful predictions. Removing the full vaccination rate from the model does not generate significant differences in model performance. Additionally, the importance of past infections diminishes as the forecasting window extends.

**Figure 5.**
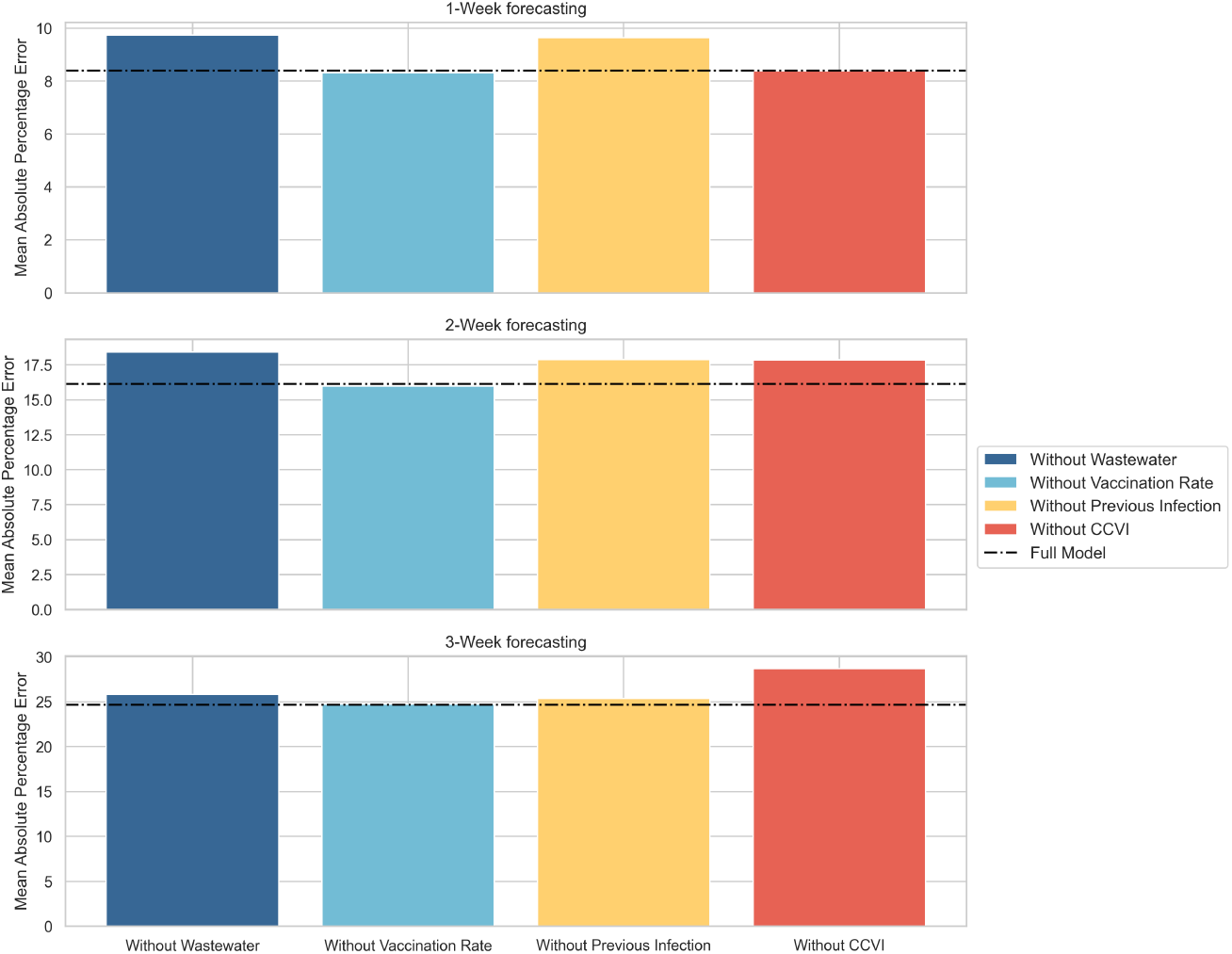
Variable contribution in model performance for 1, 2, and 3 week out continuous hospitalization forecasts. The black dotted line indicates the full model performance, and the colored bars indicate the MAPE with that variable removed. A larger increase in MAPE indicates a worse performance and a higher importance in model performance.

## 4. Discussion

### Integrating Wastewater Surveillance Data Enhances COVID-19 Hospitalization Forecasting

As traditional epidemiological datasets, such as reported cases, become less available and more subject to reporting issues over time, alternative data streams must be considered to inform short-term forecasting models. In this study, we built a comprehensive forecasting model to predict near term COVID-19 hospitalization risks in six US cities. This model incorporates novel data streams, including wastewater surveillance data and community vulnerability indices, alongside traditional epidemiological inputs such as previous infections and vaccinations. Our model performance confirms the effectiveness of this approach, particularly emphasizing the importance of wastewater surveillance data, as explored in Section 3.3.

Our variable importance analysis identified wastewater surveillance data as a critical factor in our models, particularly for forecasting city-level COVID-19 hospitalizations 1 and 2 weeks into the future (see Figure 5). Therefore, publicly available, reliable wastewater data is crucial for improving our capabilities to predict future hospitalization risks and inform both the public and public health practitioners. Wastewater disease surveillance systems, such as the CDC National Wastewater Surveillance System (NWSS), represent a centralized database that has the potential to inform disease risk assessment and bolster modeling capabilities. Thus, we encourage continued investment in and expansion of this critical surveillance tool, along with the centralized sharing of data in standardized formats.

Beyond the immediate utility of wastewater surveillance, our findings underscore the importance of local vulnerability indices on model performance. We determined that the CCVI is the most influential variable for our 3-week continuous forecasting model. This finding aligns with previous studies demonstrating the critical role of vulnerability data in predicting weekly COVID-19 hospital admissions at less-immediate forecasting windows, such as 3 and 4 weeks in the future (Li et al., 2023). The importance of CCVI in our analysis suggests that more severe health outcomes from COVID-19 infection may be amplified in vulnerable communities. As the sole static variable in the model, the community vulnerability index avoids any information decay that may arise with the temporally dependent variables, such as wastewater surveillance data. This enduring relevance underscores the need to incorporate data on both current disease dynamics, such as wastewater surveillance data, and persistent vulnerabilities, such as the CCVI, when forecasting near future infectious disease risk.

### Categorical Forecasts Support Public Health Decision Making

The well-documented challenges of forecasting traditional continuous disease outcomes during the COVID-19 pandemic (Nixon et al., 2022) motivated this study’s focus on categorical targets for hospitalization forecasting. This approach, exemplified by the Hospitalization Capacity Risk (HCR) and Hospitalization Rate Trend (HRT) categorizations defined in this study, enhances model interpretability, actionability, and robustness.

The HCR model, derived from hospitalization rates, provides hospital administrators with an intuitive understanding of capacity risk, enabling proactive resource allocation during times of high risk. Our models achieve a 75% accuracy in predicting HCR three weeks in advance, proving consistently reliable prediction across cities and forecasting windows. Exceptionally, HCR forecasts for San Francisco resulted in a uniquely high accuracy due to the city’s uniquely low hospitalization rates. While the HRT model, based on the trends of hospitalization growth rates, is a less predictable outcome, the model successfully captures 64% of weeks classified as Large Increases and 67% of weeks classified as Large Decreases for the 2-week forecasting window (Figure 4a). The focus of the HRT on directional trends, when considered in conjunction with HCR, offers decision-makers a more comprehensive understanding of potential outbreak scenarios, facilitating timely and targeted interventions.

The inherent stability of the two categorical target variables defined in this study, which are less sensitive to minor data fluctuations than continuous outputs, further reinforces the reliability of our approach and provides a robust foundation for data-driven decision-making in the face of evolving public health challenges.

### Evaluating Forecasting Performance at Change Points Illuminates Model Capabilities

A notable issue with COVID-19 forecasting during the pandemic was the inability to accurately predict inflection points in case and hospitalization trends (Lopez et al., 2024; Cramer et al., 2021). To assess the effectiveness of our categorical models in addressing this challenge, we evaluated performance at change points, examining the models’ ability to accurately account for shifts in the COVID-19 hospitalization category. As outlined in section 3.2, we evaluated model performance by determining accuracy at all change points, as well as at upward and downward shifts specifically.

The results outlined in section 3.2 presented several important insights that merit further discussion. First, we note that our HCR model generally outperforms the HRT model for all change point evaluation methods. This aligns with the overall model performance in section 3.1, where the HRT model’s weaker performance could be attributed to its prediction of highly variable hospitalization growth rates. Second, we highlight that error metrics at change points generally worsen as forecasting window increases. This is also consistent with the results in section 3.1, which suggests that this decline is likely due to the decreased correlations between wastewater and future hospitalizations over time. Third, we recognize that the performance at change points is weaker for both models compared to overall model accuracy, as per Table 5 the difference in WMSE is less than 0.3 across all models and forecast horizons. While there is still room for improvement, this indicates that the model is reasonably effective at predicting the correct category at change points. Nevertheless, this discrepancy in model performance at change points further highlights the need for a greater focus on improving forecasting accuracy during periods of rapid changes.

Our change point analysis methodology evaluates the utility of categorical forecasting models at critical points. Sudden shifts in population level disease dynamics are some of the most crucial trends to understand in order to better inform decision makers at times of great uncertainty. Our change point analysis enhances model utility by evaluating how well each categorization captures hospitalization dynamics during periods of rapid change. We believe that this straightforward yet comprehensive approach to assessing categorical model accuracy at change points is an effective and transferable method for evaluating model performance and developing timely policies to mitigate the effects of a virus.

### City-Level COVID-19 Hospitalization Forecasting is Effective and Useful

We propose that achieving a balance between forecast accuracy and actionable spatial scales, such as cities, is crucial for providing key insights that can be translated into effective mitigation strategies for COVID-19 and other infectious diseases. COVID-19 forecasting using wastewater surveillance has been implemented at several different granularities, such as country, state, county, city, and sewershed level. (Schenk et al., 2023; Hill et al., 2023; Li et al., 2023; Galani et al., 2022). We contend that forecasting at a city-level, which has received the least attention by modelers and is well-aligned with the spatial boundaries of wastewater data, is particularly advantageous for decision-makers. Densely populated urban areas are often the epicenter of COVID-19 infections and have government structures that allow for more targeted public health interventions. Thus, accurate city-level forecasts can more effectively help inform local government policies to prevent harm to the population and minimize economic burden at these disease epicenters (Lopez et al., 2024). Despite this potential, city-level forecasting is an under-utilized tool, with most forecasts targeting larger, aggregated populations such as county-level, state-level, and national-level (Sen et al., 2023). This is often due to the lack of reliable city-level disease surveillance, a challenge addressed by the natural alignment between wastewater treatment plants and city boundaries as demonstrated in Figure S1 in **Supplementary A**. Additionally, many cities have multiple wastewater treatment facilities which can be aggregated to provide a smoother prediction (Medina et al., 2022). Multiple wastewater treatment facilities also enable higher modeling granularities within cities. Overall, WBE provides an opportunity to advance higher resolution infectious disease forecasting and permits local governments to implement mitigation strategies that best suits their communities.

We note that the quality of wastewater data significantly impacts model performance across cities. Houston and New York City consistently demonstrate superior performance due to their high-quality wastewater data, characterized by consistent reporting and a large number of sewersheds. Conversely, as shown in Figures 3b and 4b, Charlotte and Denver exhibit poorer model performance for both HCR and HRT due to lesser data quality, stemming from recorded lapses in reporting or changes in reporting styles (Keshaviah et al., 2023; Ghanbari et al., 2024). This finding underscores the importance of consistent and transparent WBE data for both understanding local trends and enhancing forecasting capabilities.

### Limitations and Future Work

Wastewater disease surveillance for COVID-19 is a rapidly evolving field. Different cities and wastewater treatment plants have varying timelines for reporting and normalization techniques that make retrospective data difficult to compare to currently reported information. Despite the expansion of the NWSS to expand wastewater disease surveillance systems, a lack of data accessibility and inconsistencies in data collection, standardization and preprocessing can still impact data quality across locations. Consequently, the observed variation in model performance across cities can be attributed to these inherent data quality differences.

The models in this study are better equipped to capture downward shifts, which may be reflective of limited input data streams. Therefore, we propose including alternative disease prevalence information, such as circulating variants, which may improve model performance, as demonstrated in (Du et al., 2023). Wastewater-based epidemiology is a vehicle to understand community disease prevalence, and variant information can be easily recovered from it to strengthen model performance, something considered for future extensions of this work.

In this study, we focus on aggregating epidemiological data to a city level to create a more granular forecasting scale than the traditional state or county-level forecasts. By utilizing wastewater disease surveillance, we are able to provide an even more specific geographical scale at which to understand disease dynamics. In future work, we aim to expand our modeling capabilities to understand wastewater signals at the sewershed level as an early warning signal for smaller spatial scales.

## 5. Conclusions

In this study, we explored the use of wastewater disease surveillance data for categorical COVID-19 forecasting at the city level with two models: the Hospitalization Capacity Risk (HCR) categorization and the Hospitalization Rate Trend (HRT) categorization. The HCR model provides an understanding of general risk based on bed availability to inform resource allocation strategies. The HRT Categorization forecasts the growth rate of hospitalizations to understand the predicted trajectory of burden on a healthcare system. We also proposed a methodology to determine the circumstances under which models are useful with our change point analysis. Utilizing these models, we demonstrate that wastewater disease surveillance data is a crucial input for COVID-19 hospitalization forecasting. Due to the natural alignment of city borders and sewersheds, and the ability to generate reliable forecasts, city-level forecasting utilizing wastewater disease surveillance data is an advantageous approach. As the community dynamics of COVID-19 change and data availability shifts, we must update our forecasting capabilities to use more impactful inputs and design more beneficial and interpretable targets.

## Data Availability

All data produced are available online

https://github.com/nahomiiiie/WastewaterForecasting

## 1. Supplementary A: Methods

### 1.1 Data Collection

#### Sewersheds and City Boundaries

The sewershed maps are provided by city services for each location, the references for which can be found in the image caption. The city boundary and ZIP code maps were created using the Python package GEOPANDAS. The city boundaries are generated using data obtained from the U.S. Department of Health and Human Services (US HHS, 2023). We delineated these maps with the USPS ZIP codes, as identified by the USPS ZIP code lookup tool (United States Postal Service, 2024). To properly represent the USPS ZIP codes on the map, we overlaid the USPS ZIP codes with the ZIP Code Census Tract Areas, obtained from the United States Census Bureau (Census, 2020). Therefore, any ZIP code that did not correspond with a physical polygon (i.e., ZIP code that referred to a single building) was removed from these maps.

**Figure 1.**
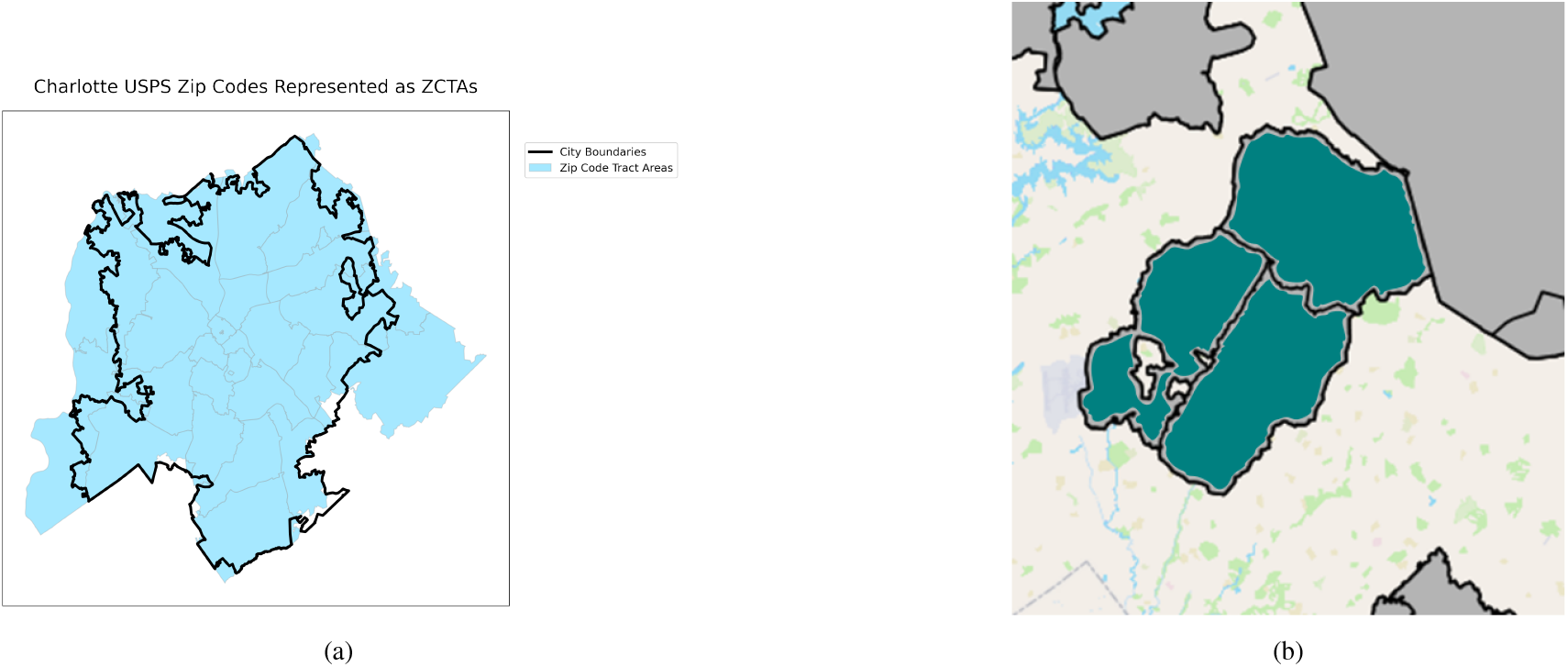
A side by side comparison of A) Charlotte city boundaries delineated by USPS Zip Code and B) the sewershed map of Charlotte. The sewersheds included in the study are highlighted in blue. (North Carolina Department of Health and Human Services, 2024)

**Figure 2.**
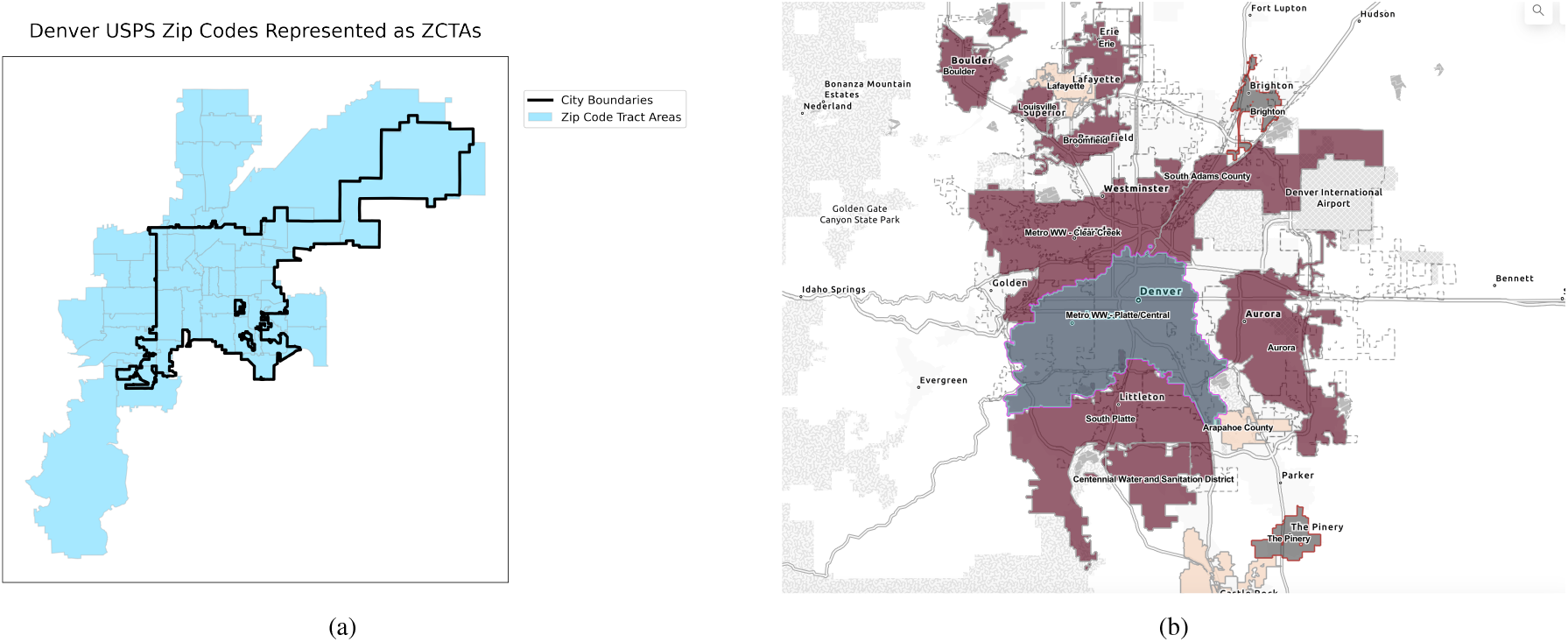
A side by side comparison of A) Denver city boundaries delineated by USPS Zip Code and B) the sewershed map of Colorado. The included sewershed is highlighted in blue. (Colorado Department of Public Health and Environment, 2024)

#### Cross Correlations Between Wastewater and Hospitalizations

In order to determine the temporal relationship between wastewater SARS-CoV-2 rates and COVID-19 hospitalizations, we examine the cross correlation between these two time series. As shown in Figure 7, the weeks when the lag is greater than 0.75 indicate that wastewater SARS-CoV-2 rates are highly correlated with hospitalizations that many weeks in advance. For example, in Houston, TX, the wastewater SARS-CoV-2 rates are highly correlated for the observed week of hospitalizations, as well as the preceding 35 weeks. We included the SARS-CoV-2 rates from the previous three weeks in the model as predictor variables to align with the most common correlations observed across all cities.

**Figure 3.**
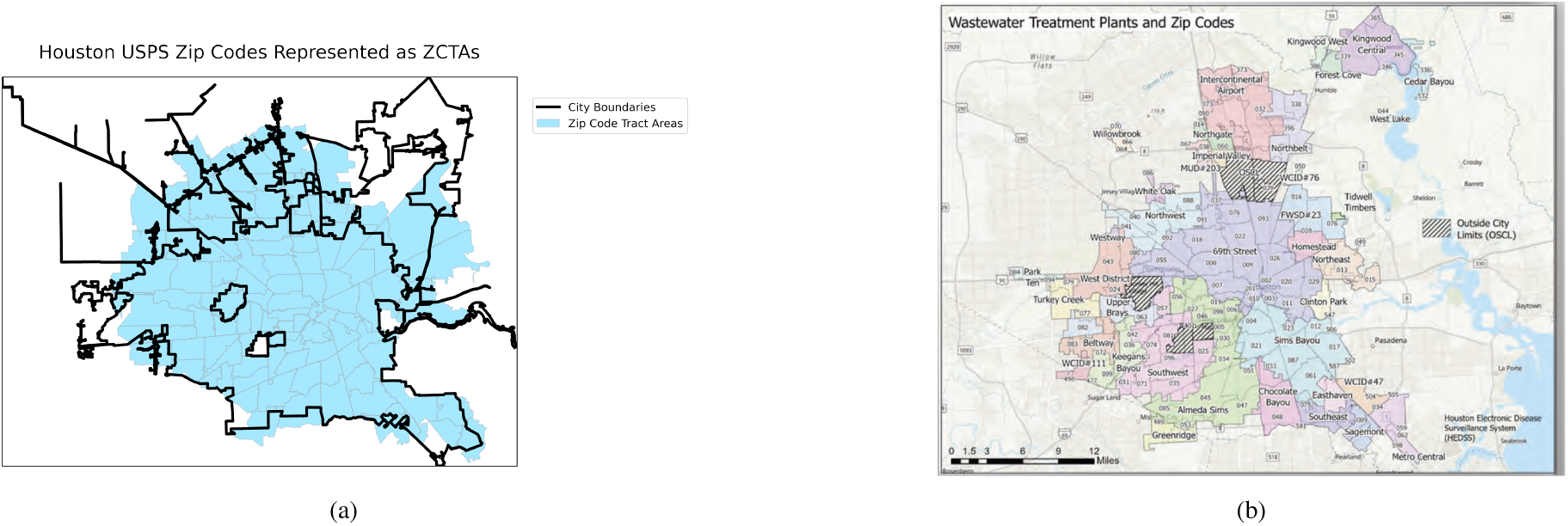
A side by side comparison of A) Houston city boundaries delineated by USPS Zip Code and B) the sewershed map of Houston (Hopkins et al., 2022)

**Figure 4.**
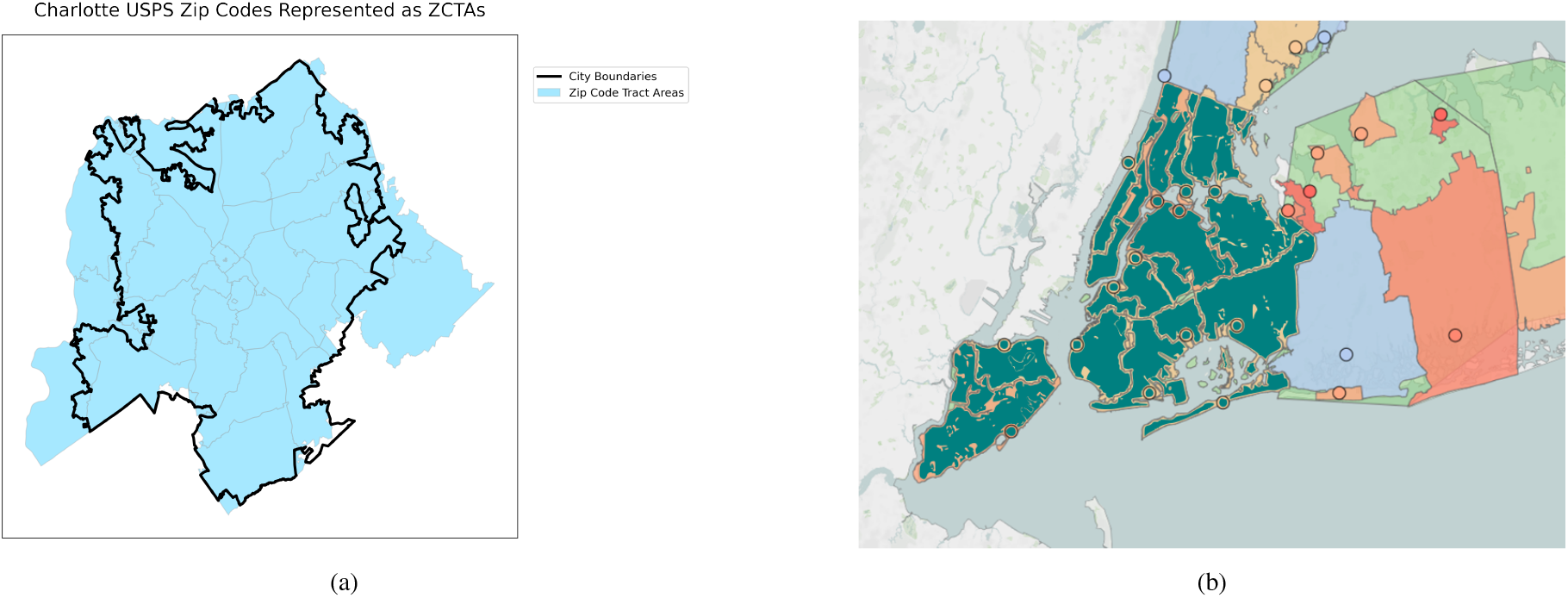
A side by side comparison of A) New York City boundaries delineated by USPS Zip Code and B) the sewershed map of New York City CITATION

**Figure 5.**
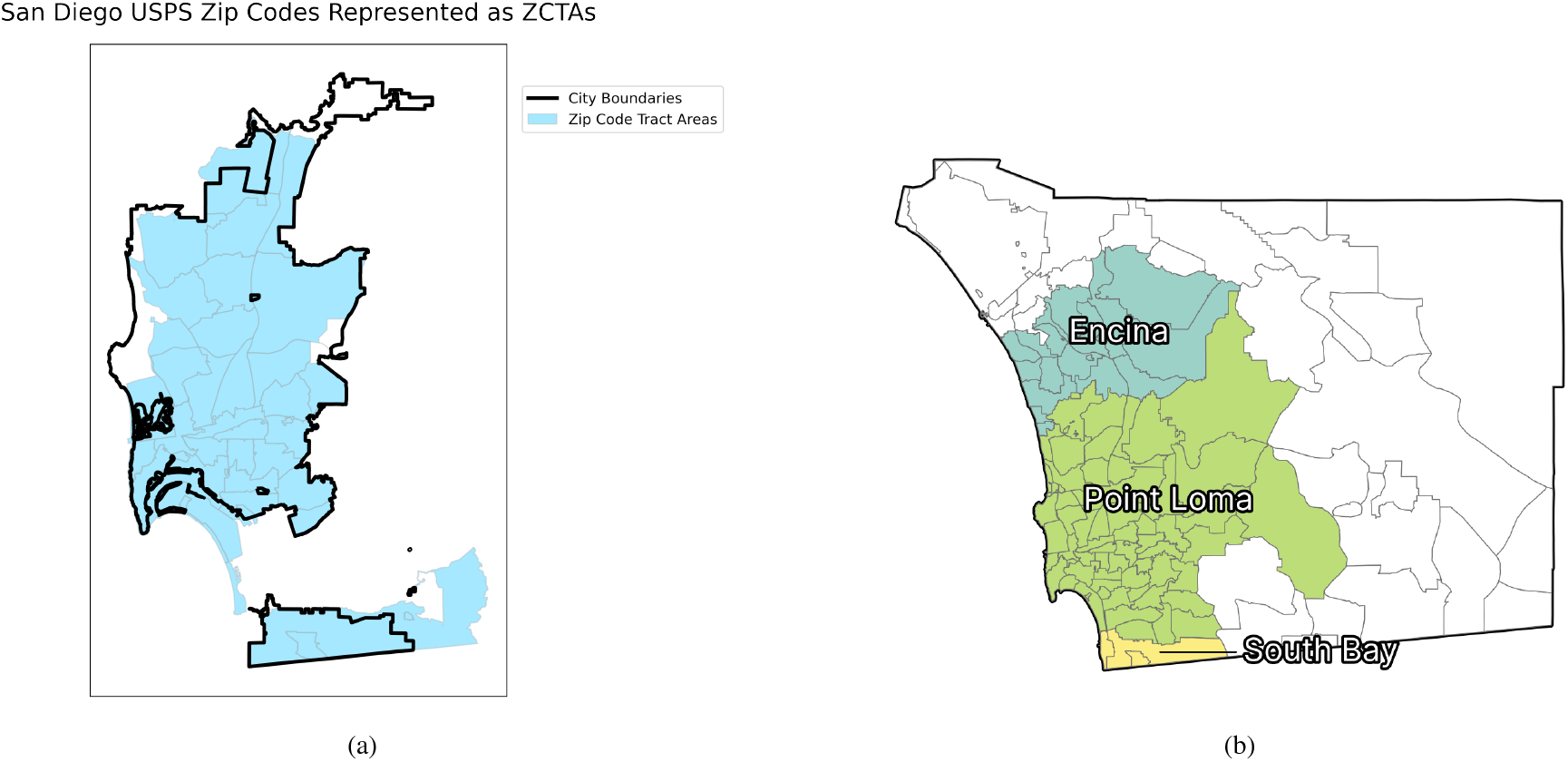
A side by side comparison of A) San Diego city boundaries delineated by USPS Zip Code and B) the sewershed map of San Diego: (Lab, 2024)

**Figure 6.**
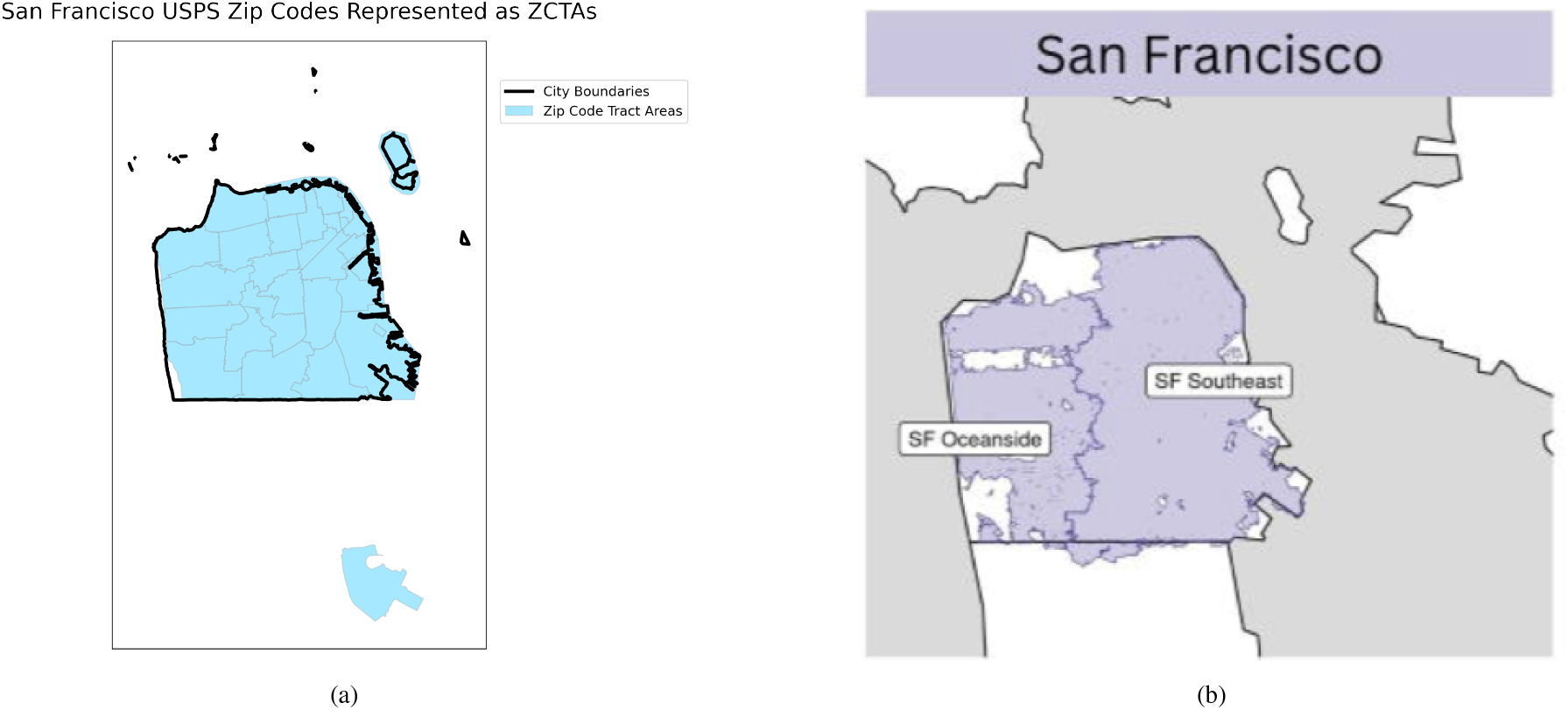
A side by side comparison of A) San Francisco city boundaries delineated by USPS Zip Code and B) the sewershed map of San Francisco: (Ravuri et al., 2024)

### 1.2 Input Variables

### 1.3 Target Design

The 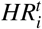 is obtained from the U.S. Department of Health and Human Services (HHS) COVID-19 Reported Patient Impact and Hospital Capacity by Facility dataset at HealthData.gov (United States Government Department of Health & Human Services, 2024). This dataset has weekly aggregated facility-level data for hospital utilization, derived from reports from HHS TeleTracking and reports made directly to HHS Protect from state and territorial health departments. The number of hospitalizations for each facility can be derived from the reported 7-day sum of confirmed and suspected adult COVID-19 cases, which details the total number of patients counted at that facility that week, and the 7-day coverage, which details the number of days that the number of patients were recorded at each facility that week. In the dataset, if the reported 7-day sum was less than 4, the cell was replaced with a -999999 to protect patient confidentiality. For the indicated low values, we interpolate the number of hospitalizations that week as the mean value of the surrounding weeks. If there were multiple missing values in a row, we replace the missing sum with a 2, as the average value between the 1 and 3 patients that the missing value represents. Using these reported and interpolated values, we calculate the average number of patients seen that week as the 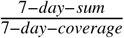 for all facilities.

### 1.4 Error Metrics

In this section we provide the equations that define our five error metrics: 1) Accuracy, 2) Mean Square Error (MSE), 3) Weighted Mean Square Error (WMSE), 4) Brier Score, and 5) Brier Skill Score. To calculate the error metrics, we convert our forecasted categories to numerical values; For the HRT, the labels of [Large Decrease, Decrease, Stable, Increase, and Large Increase] map to [-2, -1, 0, 1, 2]. For the HCR, the labels of [Very Low, Low, Moderate, High, Very High] map to [1, 2, 3, 4, 5].

Accuracy measures the percent of predicted labels that match the true labels, and is defined as

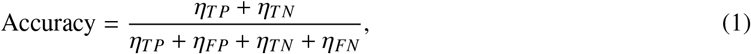

where *η*_*TP*_, *η*_*FP*_, *η*_*TN*_, *η*_*FN*_ indicate true positives, false positives, true negatives and false negatives respectively.

MSE demonstrates the magnitude of error and is defined as

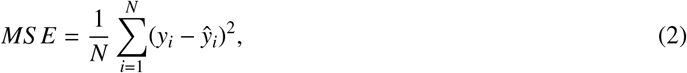

where *N* is the size of the test set, *y*_*i*_ indicates the actual category label, and ŷ_*i*_ is the predicted category.

The WMSE weight the magnitude of the error, where the weights are the probabilities of each category.

**Figure 7.**
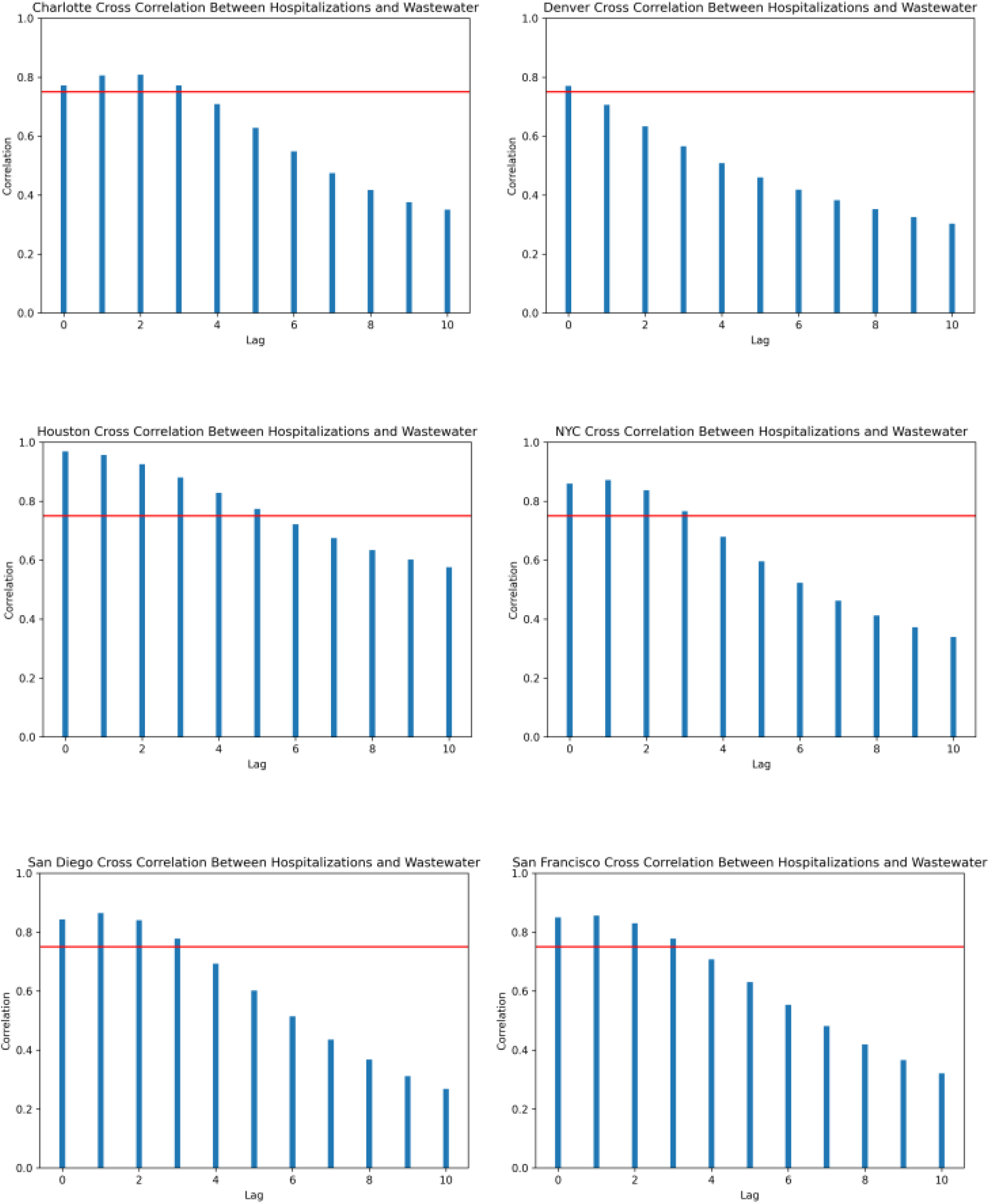
**Figure S2:** Cross correlations between wastewater SARS-CoV-2 rates and hospitalizations per 100,000 people in each city included in the study from January 2021 to September 2022. The red line indicates the critical threshold of 0.75 we consider for inclusion as a predictor variable.

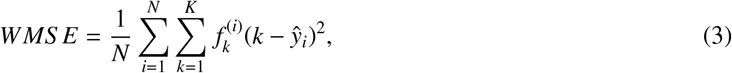

where *K* denotes the set of categories, 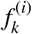 indicates the probability of *k*-th category being forecasted for the *i*-th item in the test set.

The Brier Score measures the accuracy of probabilistic forecasts. It is calculated as the mean square difference between the predicted probability of each category to the actual probability of each category. The Brier Score is defined as

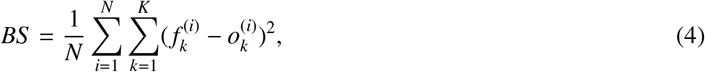

where 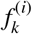 indicates the probability of *k*-th category being forecasted for the *i*-th item in the test set and 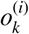 is a binary variable which is equal to 1 for the true category *k*_*i*_ and a 0 for all other categories for the *i*-th item in the test set.

The Brier Skill Score, compares model performance that of a random guess, where there is a uniform probability for each category, defined as

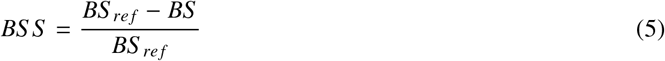

Where *BS* is the calculated Brier Score from our test set, and *BS* _*ref*_ is the Brier Score from of a random guess on the test set. A BSS less than 0 indicates that our predictions perform worse than the baseline, equal to 0 indicates that the score is equivalent to the baseline, and greater than one indicates that the our predictions perform better than the baseline.

## 1 Supplementary B: Results

### 1.1 Model Performance

This section describes the model performance for both the HCR and HRT over the 1- and 3-week forecasting window.

#### 1.1.1 HCR

HCR 1-week forecast

**Figure 1.**
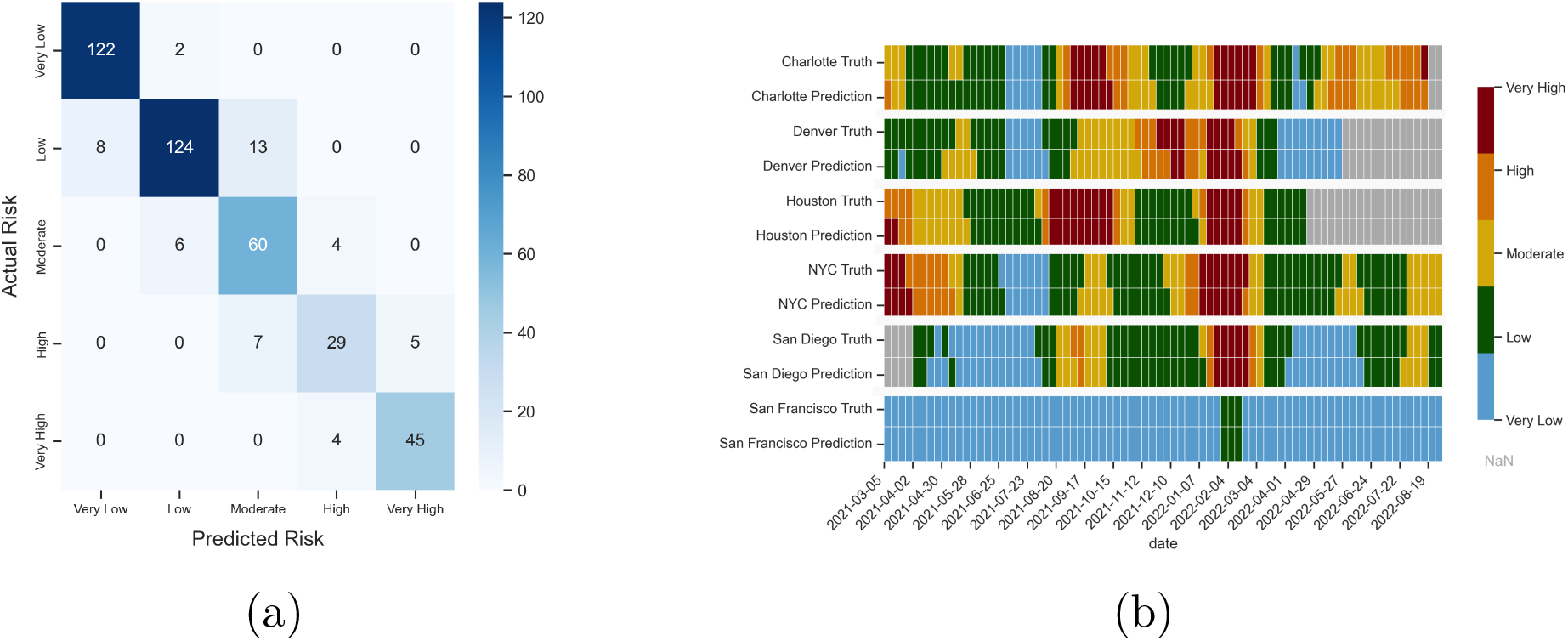
Figure S1: Model performance of Hospitalization Capacity Risk (HCR) Categorization for the 1 Week forecast. (a) Confusion Matrix detailing the accuracy of categorization for the HCR at a 1 week fore-casting window. Darker colors indicate more assignment-true label combos lie in that category. (b) Hospital Capacity Risk true and predicted label for 1-week forecasts by city for January 2021 to September 2022. Color indicates predicted category. Grey squares indicate that a prediction was not made for that week due to a lack of available data.

**Figure 2.**
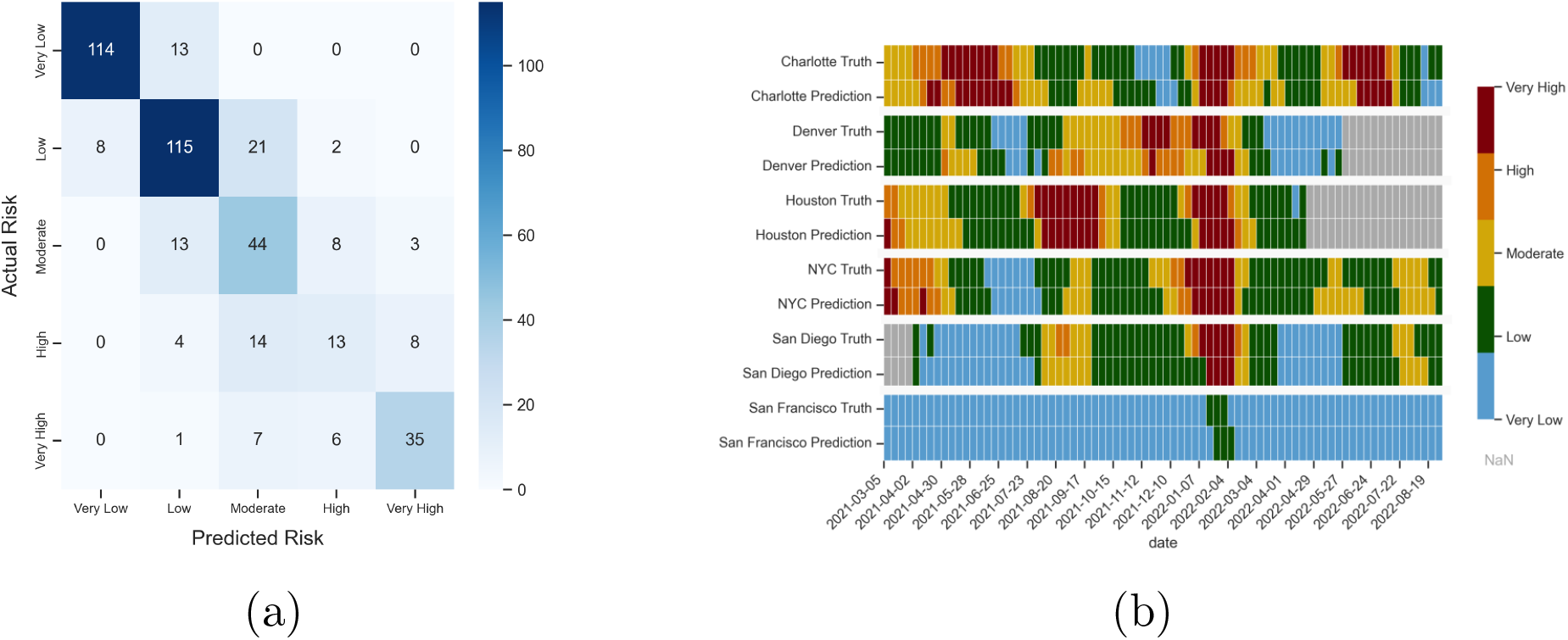
Figure S2: Model performance of Hospitalization Capacity Risk (HCR) Categorization for the 3-week forecast. (a) Confusion Matrix detailing the accuracy of categorization for the HCR over a 3 week forecasting window. Darker colors indicate more assignment-true label combos lie in that category. (b) Hospital Capacity Risk true and predicted label for 3-week forecasts by city for January 2021 to September 2022. Color indicates predicted category. Grey squares indicate that a prediction was not made for that week due to a lack of available data.

#### 1.1.2 HRT

HRT 1-week forecast

**Figure 3.**
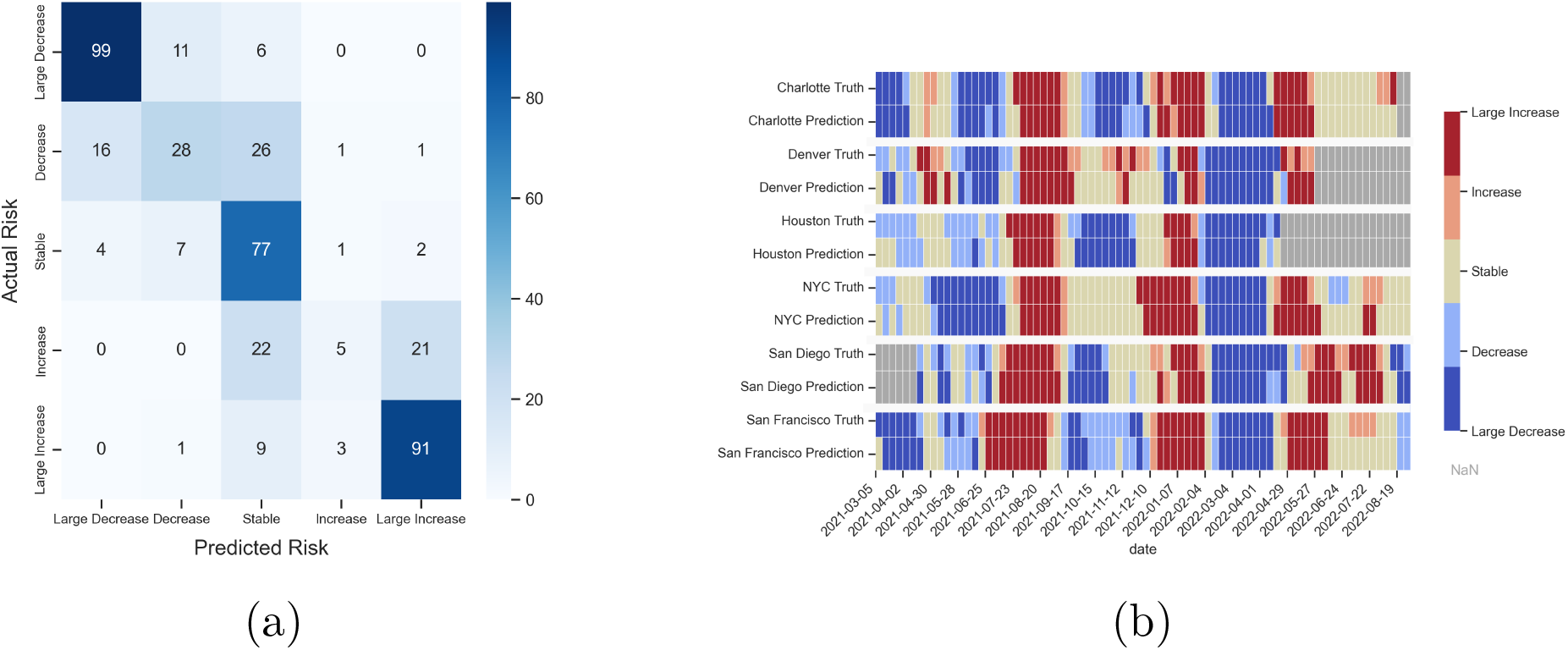
Figure S3: Model performance of Hospitalization Rate Trend (HRT) Categorization for the 1 week forecast. (a) Confusion matrix detailing the accuracy of the categorization of the HRT at a 1 week forecasting window. Darker colors indicate more assignment-true label combos lie in that category. (b) City-specific Rate Trend target and 1-week prediction over time from January 2021 to September 2022. Grey squares indicate that a prediction was not made for that week due to a lack of available data.

HRT 3-week forecast

**Figure 4.**
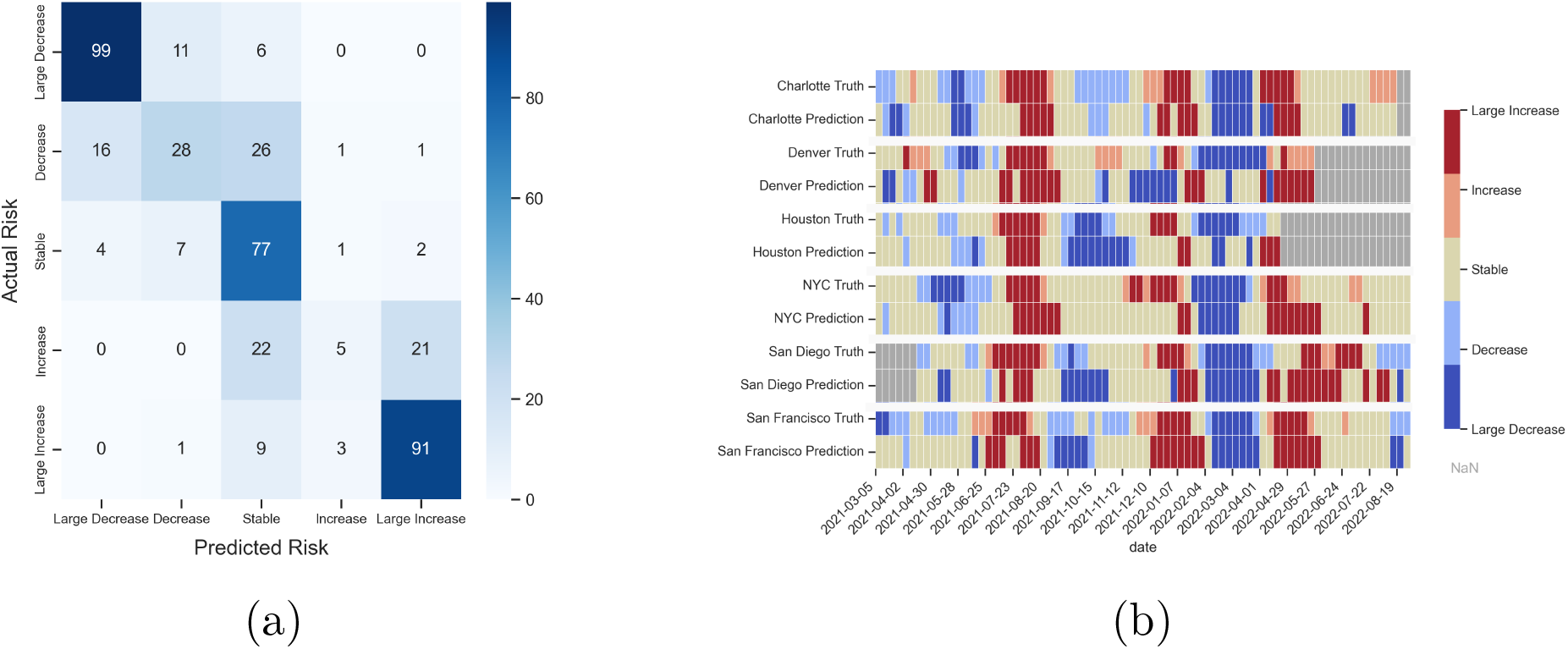
Figure S4: Model performance of Hospitalization Rate Trend (HRT) Categorization. (a) Confusion matrix detailing the accuracy of the categorization of the HRT at a 3 week forecasting window. Darker colors indicate more assignment-true label combos lie in that category. (b) City-specific Rate Trend target and 1-week prediction over time from January 2021 to September 2022. Grey squares indicate that a prediction was not made for that week due to a lack of available data.

### 1.2 Change Point Performance

**Table 1:** A summary of model performance for all change points. ↑*/*↓ denotes if an upward or downward shift change point demonstrates better performance.

|  |  | All Change Points |  |  |  |
| --- | --- | --- | --- | --- | --- |
|  |  | MSE ↓ | WMSE ↓ | BS ↓ | BSS |
| HCR | 1 Week | 0.345 | 0.244 | 0.464 | 0.420 |
|  | 2 Week | 0.614 | 0.325 | 0.703 | 0.121 |
|  | 3 Week | 1.034 | 0.454 | 0.821 | -0.027 |
| HRT | 1 Week | 0.944 | 0.628 | 0.707 | 0.117 |
|  | 2 Week | 2.304 | 1.262 | 0.881 | -0.101 |
|  | 3 Week | 2.076 | 1.100 | 0.909 | -0.136 |

**Table 2:** A summary of model performance for forecasting change points from a lower to higher category (upward shift) and a higher to lower category (downward shift). ↑*/* ↓ denotes if an upward or downward shift change point demonstrates better performance.

|  |  | Upward Shift |  |  |  | Downward Shift |  |  |  |
| --- | --- | --- | --- | --- | --- | --- | --- | --- | --- |
|  |  | MSE ↓ | WMSE ↓ | BS ↓ | BSS ↑ | MSE ↓ | WMSE ↓ | BS ↓ | BSS ↑ |
| HCR | 1 Week | 0.405 | 0.224 | 0.554 | 0.308 | 0.289 | 0.262 | 0.380 | 0.525 |
|  | 2 Week | 0.786 | 0.288 | 0.845 | -0.056 | 0.457 | 0.359 | 0.574 | 0.282 |
|  | 3 Week | 1.419 | 0.446 | 0.903 | -0.128 | 0.674 | 0.462 | 0.746 | 0.068 |
| HRT | 1 Week | 1.195 | 0.651 | 0.694 | 0.133 | 0.711 | 0.607 | 0.719 | 0.102 |
|  | 2 Week | 2.571 | 1.183 | 0.907 | -0.134 | 2.060 | 1.335 | 0.857 | -0.071 |
|  | 3 Week | 2.181 | 1.131 | 0.846 | -0.057 | 1.973 | 1.069 | 0.971 | -0.213 |

## References

Bibby, K., Bivins, A., Wu, Z., North, D., 2021. Making waves: plausible lead time for wastewater based epidemiology as an early warning system for covid-19. Water Research 202, 117438.

Bradley, A.A., Schwartz, S.S., Hashino, T., 2008. Sampling uncertainty and confidence intervals for the brier score and brier skill score. Weather and Forecasting 23, 992–1006.

California State Water Resources Control Board, 2024. Sewershed surveillance for covid-19 updates. URL: https://www.waterboards.ca.gov/resources/. accessed: 2024-07-10.

Centers for Disease Control and Prevention, 2023. Vaccines for COVID-19. https://www.cdc.gov/coronavirus/2019-ncov/vaccines/.

Centers for Disease Control and Prevention, 2024a. Cdc covid data tracker. URL: https://covid.cdc.gov/covid-data-tracker/. accessed: 2024-07-10.

Centers for Disease Control and Prevention, 2024b. Indicators for monitoring covid-19 community levels. URL: https://archive.cdc.gov/. accessed: 2024-07-10.

Colorado Department of Public Health and Environment, 2024. CDPHE COVID19 Wastewater Dashboard Data . https://data-cdphe.opendata.arcgis.com/datasets.

Cramer, E.Y., Huang, Y., Wang, Y., Ray, E.L., Cornell, M., Bracher, J., Brennen, A., Castro Rivadeneira, A.J., Gerding, A., House, K., Jayawardena, D., Kanji, A.H., Khandelwal, A., Le, K., Niemi, J., Stark, A., Shah, A., Wattanachit, N., Zorn, M.W., Reich, N.G., Consortium, U.C.F.H., 2021. The united states covid-19 forecast hub dataset. medRxiv URL: https://www.medrxiv.org/content/10.1101/2021.11.04.21265886v1, doi:10.1101/2021.11.04.21265886.

Dong, E., Du, H., Gardner, L., 2020. An interactive web-based dashboard to track covid-19 in real time. The Lancet infectious diseases 20, 533–534.

Du, H., Dong, E., Badr, H.S., Petrone, M.E., Grubaugh, N.D., Gardner, L.M., 2023. Incorporating variant frequencies data into short-term forecasting for covid-19 cases and deaths in the usa: A deep learning approach. Ebiomedicine 89.

Du, H., Saiyed, S., Gardner, L.M., 2024a. Association between vaccination rates and covid-19 health outcomes in the united states: a population-level statistical analysis. BMC Public Health 24, 220.

Du, H., Zhao, J., Zhao, Y., Xu, S., Lin, X., Chen, Y., Gardner, L.M., Yang, H.F., 2024b. Advancing real-time pandemic forecasting using large language models: A covid-19 case study arXiv:2404.06962.

Feng, S., Roguet, A., McClary-Gutierrez, J.S., Newton, R.J., Kloczko, N., Meiman, J.G., McLellan, S.L., 2021. Evaluation of sampling, analysis, and normalization methods for sars-cov-2 concentrations in wastewater to assess covid-19 burdens in wisconsin communities. Acs Es&T Water 1, 1955–1965.

FluSight-forecast-hub, 2024. FluSight 2023-2024. https://github.com/cdcepi/FluSight-forecast-hub/tree/main.

Galani, A., Aalizadeh, R., Kostakis, M., Markou, A., Alygizakis, N., Lytras, T., Adamopoulos, P.G., Peccia, J., Thompson, D.C., Kontou, A., et al., 2022. Sars-cov-2 wastewater surveillance data can predict hospitalizations and icu admissions. Science of The Total Environment 804, 150151.

Ghanbari, M., Huang, J., Luc, A., Arabi, M., Goldman, J.E., Byrne-Nash, R., Kane, S.J., Ferrell, R., Fielder, T., De Long, S.K., Wilusz, C.J., 2024. View of an evolving pandemic: Changes in the relationship between clinical cases and levels of sars-cov-2 rna in colorado wastewater. ACS ES&T Water 4, 2018–2030. URL: 10.1021/acsestwater.3c00615, doi:10.1021/acsestwater.3c00615, arXiv:10.1021/acsestwater.3c00615.

Hill, D.T., Alazawi, M.A., Moran, E.J., Bennett, L.J., Bradley, I., Collins, M.B., Gobler, C.J., Green, H., Insaf, T.Z., Kmush, B., et al., 2023. Wastewater surveillance provides 10-days forecasting of covid-19 hospitalizations superior to cases and test positivity: A prediction study. Infectious disease modelling 8, 1138–1150.

Hillary, L.S., Malham, S.K., McDonald, J.E., Jones, D.L., 2020. Wastewater and public health: the potential of wastewater surveillance for monitoring covid-19. Current Opinion in Environmental Science & Health 17, 14–20.

Kanchan, S., Ogden, E., Kesheri, M., Skinner, A., Miliken, E., Lyman, D., Armstrong, J., Sciglitano, L., Hampikian, G., 2024. Covid-19 hospitalizations and deaths predicted by sars-cov-2 levels in boise, idaho wastewater. Science of The Total Environment 907, 167742.

Keshaviah, A., Diamond, M.B., Wade, M.J., Scarpino, S.V., Ahmed, W., Amman, F., Aruna, O., Badilla-Aguilar, A., Bar-Or, I., Bergthaler, A., et al., 2023. Wastewater monitoring can anchor global disease surveillance systems. The Lancet Global Health 11, e976–e981.

Kinder Institute Urban Data Platform, 2024. Dataset catalog. URL: https://www.kinderudp.org. accessed: 2024-07-10.

Lab, A., 2024. Sars-cov-2 wastewater san diego. URL: https://github.com/andersen-lab. accessed: 2024-07-10.

Li, X., Liu, H., Gao, L., Sherchan, S.P., Zhou, T., Khan, S.J., Van Loosdrecht, M.C., Wang, Q., 2023. Wastewater-based epidemiology predicts covid-19-induced weekly new hospital admissions in over 150 usa counties. Nature Communications 14, 4548.

Lopez, V.K., Cramer, E.Y., Pagano, R., Drake, J.M., O’Dea, E.B., Adee, M., Ayer, T., Chhatwal, J., Dalgic, O.O., Ladd, M.A., Linas, B.P., Mueller, P.P., Xiao, J., Bracher, J., Castro Rivadeneira, A.J., Gerding, A., Gneiting, T., Huang, Y., Jayawardena, D., Kanji, A.H., Le, K., Mühlemann, A., Niemi, J., Ray, E.L., Stark, A., Wang, Y., Wattanachit, N., Zorn, M.W., Pei, S., Shaman, J., Yamana, T.K., Tarasewicz, S.R., Wilson, D.J., Baccam, S., Gurung, H., Stage, S., Suchoski, B., Gao, L., Gu, Z., Kim, M., Li, X., Wang, G., Wang, L., Wang, Y., Yu, S., Gardner, L., Jindal, S., Marshall, M., Nixon, K., Dent, J., Hill, A.L., Kaminsky, J., Lee, E.C., Lemaitre, J.C., Lessler, J., Smith, C.P., Truelove, S., Kinsey, M., Mullany, L.C., Rainwater-Lovett, K., Shin, L., Tallaksen, K., Wilson, S., Karlen, D., Castro, L., Fairchild, G., Michaud, I., Osthus, D., Bian, J., Cao, W., Gao, Z., Lavista Ferres, J., Li, C., Liu, T.Y., Xie, X., Zhang, S., Zheng, S., Chinazzi, M., Davis, J.T., Mu, K., Pastorey Piontti, A., Vespignani, A., Xiong, X., Walraven, R., Chen, J., Gu, Q., Wang, L., Xu, P., Zhang, W., Zou, D., Gibson, G.C., Sheldon, D., Srivastava, A., Adiga, A., Hurt, B., Kaur, G., Lewis, B., Marathe, M., Peddireddy, A.S., Porebski, P., Venkatramanan, S., Wang, L., Prasad, P.V., Walker, J.W., Webber, A.E., Slayton, R.B., Biggerstaff, M., Reich, N.G., Johansson, M.A., 2024. Challenges of covid-19 case forecasting in the us, 2020–2021. PLOS Computational Biology 20, 1–25. URL: 10.1371/journal.pcbi.1011200, doi:10.1371/journal.pcbi.1011200.

Medina, C.Y., Kadonsky, K.F., Roman Jr, F.A., Tariqi, A.Q., Sinclair, R.G., D’Aoust, P.M., Delatolla, R., Bischel, H.N., Naughton, C.C., 2022. The need of an environmental justice approach for wastewater based epidemiology for rural and disadvantaged communities: a review in california. Current Opinion in Environmental Science & Health 27, 100348.

New York City Department of Health and Mental Hygiene, 2024. Sars-cov-2 concentrations measured in nyc wastewater. URL: https://data.cityofnewyork.us/Health/SARS-CoV-2-concentrations-measured-in-NYC-Wastewat. accessed: 2024-07-10.

Nixon, K., Jindal, S., Parker, F., Marshall, M., Reich, N.G., Ghobadi, K., Lee, E.C., Truelove, S., Gardner, L., 2022. Real-time covid-19 forecasting: challenges and opportunities of model performance and translation. The Lancet Digital Health 4, e699–e701.

North Carolina Department of Health and Human Services, 2024. NC COVID-19 Dashboard Data . https://covid19.ncdhhs.gov/dashboard/data-behind-dashboards.

Pooley, N., Abdool Karim, S.S., Combadière, B., Ooi, E.E., Harris, R.C., El Guerche Seblain, C., Kisomi, M., Shaikh, N., 2023. Durability of vaccine-induced and natural immunity against covid-19: a narrative review. Infectious Diseases and Therapy 12, 367–387.

Reese, H., Iuliano, A.D., Patel, N.N., Garg, S., Kim, L., Silk, B.J., Hall, A.J., Fry, A., Reed, C., 2021. Estimated incidence of coronavirus disease 2019 (covid-19) illness and hospitalization—united states, february–september 2020. Clinical Infectious Diseases 72, e1010–e1017.

Rubin, R., 2021. Covid-19 testing moves out of the clinic and into the home. Jama 326, 1362–1364.

Schenk, H., Heidinger, P., Insam, H., Kreuzinger, N., Markt, R., Nägele, F., Oberacher, H., Scheffknecht, C., Steinlechner, M., Vogl, G., et al., 2023. Prediction of hospitalisations based on wastewater-based sars-cov-2 epidemiology. Science of The Total Environment 873, 162149.

Sen, A., Stevens, N.T., Tran, N.K., Agarwal, R.R., Zhang, Q., Dubin, J.A., 2023. Forecasting daily covid-19 cases with gradient boosted regression trees and other methods: evidence from us cities. Frontiers in Public Health 11, 1259410.

Shah, S., Gwee, S.X.W., Ng, J.Q.X., Lau, N., Koh, J., Pang, J., 2022. Wastewater surveillance to infer covid-19 transmission: A systematic review. Science of The Total Environment 804, 150060.

United States Government Department of Health & Human Services, 2024. COVID-19 Reported Patient Impact and Hospital Capacity by Facility. https://healthdata.gov/d/t7zc-4t6g.

United States Postal Service, 2024. Zip code lookup. URL: https://tools.usps.com/zip-code-lookup.htm. accessed: 2024-07-10.

Vaughan, L., Zhang, M., Gu, H., Rose, J.B., Naughton, C.C., Medema, G., Allan, V., Roiko, A., Blackall, L., Zamyadi, A., 2023. An exploration of challenges associated with machine learning for time series forecasting of covid-19 community spread using wastewater-based epidemiological data. Science of The Total Environment 858, 159748.

Ventures, S., 2021. Vulnerable communities and covid-19: the damage done, and the way forward. Washington, DC: Surgo Ventures .

Yang, H., Sürer, Ö., Duque, D., Morton, D.P., Singh, B., Fox, S.J., Pasco, R., Pierce, K., Rathouz, P., Valencia, V., et al., 2021. Design of covid-19 staged alert systems to ensure healthcare capacity with minimal closures. Nature communications 12, 3767.

Zamarreño, J.M., Torres-Franco, A.F., Gonçalves, J., Muñoz, R., Rodríguez, E., Eiros, J.M., García-Encina, P., 2024. Wastewater-based epidemiology for covid-19 using dynamic artificial neural networks. Science of The Total Environment 917, 170367.

## References

Census, 2020. Census zip code tabulation areas. URL: https://www.census.gov/cgi-bin/geo/shapefiles. accessed: 2024-07-30.

Hopkins, L., Ensor, K., Stadler, L.B., 2022. Best practices for wastewater-based epidemiology URL: https://www.hou-wastewater-epi.org/sites/g/files/bxs4786/files/2022-06/COH-Wastewater-Epi-Best-Practices.pdf. accessed: 21 June 2024.

Ravuri, S., Burnor, E., Routledge, I., Linton, N., Thakur, M., Boehm, A., Wolfe, M., Bischel, H.N., Naughton, C.C., Yu, A.T., White, L.A., Leoón, T.M., 2024. “real-time county-aggregated wastewater-based estimates for sars-cov-2 effective reproduction numbers”. medRxiv URL: https://www.medrxiv.org/content/early/2024/05/03/2024.05.02.24306456, doi:10.1101/2024.05.02.24306456, arXiv:https://www.medrxiv.org/content/early/2024/05/03/2024.05.02.24306456.full.pdf.

US HHS, 2023. 500 cities: City boundaries. URL: https://catalog.data.gov/dataset/500-cities-city-boundaries. accessed: 2024-07-30.

